# Time-Aware Contrastive Transformer for Longitudinal Patient Representation Learning

**DOI:** 10.64898/2026.06.23.26356236

**Authors:** Anni Heinolainen, Linh Tran, Arushi Arora, Suvi Silén, Risto Renkonen, Miika Koskinen

**Author notes:** Corresponding author: Anni Heinolainen.

## Abstract

Learning high-quality longitudinal patient representations from irregular electronic health records (EHRs) is essential for understanding heterogeneity in time-evolving diseases such as cancer. Longitudinal patient representation learning methods often rely on external labels for downstream tasks or do not model the temporal dynamics between medical events explicitly, reducing the clinical applicability of learned disease trajectories. In this work, we propose the Time-Aware-Contrastive-Transformer (TACT), a transformer-based model that integrates explicit temporal modeling with a fully self-supervised contrastive learning framework. We introduce a sampling-based data augmentation workflow that leverages hierarchical taxonomies of diagnoses and medications to enrich representation learning. Evaluated on a large real-world dataset, TACT demonstrates robust performance across patient representation and event embedding metrics and outperforms two time-aware transformer comparison models. Unlike the comparison models, TACT successfully bridges contrastive learning with medical hierarchies, allowing it to track precise disease trajectories and discover clinically actionable patient phenotypes. Consequently, this approach establishes a comprehensive framework for characterizing patient heterogeneity through the identification of potentially clinically meaningful subgroups with distinct progression profiles.

## 1 Introduction

Longitudinal electronic health records (EHRs) are a rich but underutilized data source for characterizing disease progression patterns and their heterogeneity within patient populations. Learning high-quality, generalizable patient representation vectors is essential for capturing this heterogeneity, particularly in complex time-evolving diseases such as cancer. These representations may reveal clinically meaningful patient subgroups with distinct disease progression patterns. However, current approaches in representation learning with EHRs focus mainly on regular-time data [1] or rely on external labels for downstream tasks, which can cause the learned representation space to collapse around the labels [2], thus limiting their ability to generalize beyond the specified downstream task [3].

The longitudinal and irregular nature of medical event sequences poses a significant challenge for modeling. Traditional approaches such as imputation [4], time discretization [5], and modeling short time windows [6] may be insufficient for long event sequences. Recently, transformer-based models with time-aware components have been introduced to address both of these challenges [7],[8]. Nevertheless, even transformers typically rely on labeled downstream objectives during fine-tuning [9]. Universally accepted labels for characterizing disease progression across clinical settings are lacking, highlighting the need for self-supervised methods, such as contrastive learning [10].

Contrastive learning is a self-supervised framework for learning discriminative representations without relying on external labels for downstream tasks. In this approach, the unsupervised learning problem is formulated as a classification problem: Each sample is augmented to create an original-augmented pair, which is treated as a positive pair during loss calculation, while all other samples and their augmentations are considered negative pairs. The model is trained using contrastive loss function, which is designed to pull the representations of positive pairs closer in the latent space, while pushing apart the negative pairs. [2]

In the medical setting, contrastive learning has been adapted for both text and structured data [11],[12]. For structured EHRs, augmentations typically include adding noise to numerical data [13], permuting the order of events within a data modality [14], random deletion, and synonym insertion [12]. Diagnoses and medications are key components of EHR data, and their coding systems - International Statistical Classification of Diseases and Related Health Problems, 10th Revision (ICD-10) [15] for diagnoses and Anatomical Therapeutic Chemical (ATC) [16] for medications - are inherently hierarchical. Incorporating this hierarchy allows for more precise encoding of clinical relationships in event embeddings. While the hierarchy of diagnosis codes has been incorporated into augmentation workflows [17],[18], similar hierarchical approaches have not yet been applied to ATC medication codes in this context.

Recent work has successfully combined transformers with contrastive learning [19],[20]. However, existing frameworks focus on multi-source data fusion [19], evaluate learned representations on downstream tasks [21], neglect data modality [12], or overlook temporal relationships [13]. Transformers with explicit modeling of both data modality and time-awareness have not yet been applied within a fully self-supervised contrastive learning framework. We hypothesize that incorporating modality information will improve event embedding quality and enable more accurate identification of disease progression patterns.

To address these limitations, we propose the Time-Aware Contrastive Transformer (TACT), which combines the time-aware transformer ClinicalTAAT [8] with a contrastive learning framework and hierarchical pre-training modules for embedding diagnosis and medication codes. TACT learns generalizable patient representations that capture interactions between medical events and their timings in a fully self-supervised manner. Our contributions are: (i) a hierarchical, sampling-based augmentation protocol for structured EHRs; (ii) a transformer-based, self-supervised contrastive learning model (TACT) for learning high-quality patient representations from irregular and sparse EHR data; and (iii) a rigorous evaluation of the strengths and limitations of TACT relative to typical masked pre-training protocol.

## 2 Methods

The workflow of the experiments is shown in Figure 1 and repeatedly occurring variables in Table 1

**Table 1.** Summary of mathematical variables used in the method section.

| Variable | Definition |
| --- | --- |
| $N$ | Number of patients |
| $M$ | Number of observed events |
| $K$ | Number of classes |
| $n_b$ | Number of patients in batch |
| $n_e$ | Number of unique events in the dataset |
| $n_c$ | Number of event embeddings in class $c$ |
| $d_e$ | Embedding dimension |
| $\max_l$ | Maximum number of events in an event sequence across all included event sequences |
| $\mathbf{Z} \in \mathbb{R}^{N \times d_e}$ | Patient representations |
| $\mathbf{Z}_{\text{sequence}} \in \mathbb{R}^{N \times \max_l \times d_e}$ | Contextualized event sequence representations |
| $\mathbf{z} \in \mathbb{R}^{d_e}$ | Representation of one patient |
| $\tilde{\mathbf{z}} \in \mathbb{R}^{d_e}$ | Representation of augmentation |
| $\text{Sc}(\cdot, \cdot)$ | Cosine similarity function |
| $\mathcal{E}_c$ | Set of event embeddings belonging to class $c$ |
| $\mathbf{e}^{\text{category}} \in \mathbb{R}^{d_e}$ | Embedding of event category |
| $\mathbf{e}^{\text{name}} \in \mathbb{R}^{d_e}$ | Embedding of event name |
| $\mathbf{e}^{\text{value}} \in \mathbb{R}^{d_e}$ | Embedding of event value |
| $\mathbf{e}^{\text{combined}} = \mathbf{e}^{\text{category}} + \mathbf{e}^{\text{name}}$ | Combined event embedding |
| $\mathbb{I}(\cdot)$ | Indicator function, takes values $\{0, 1\}$ |

**Figure 1.**
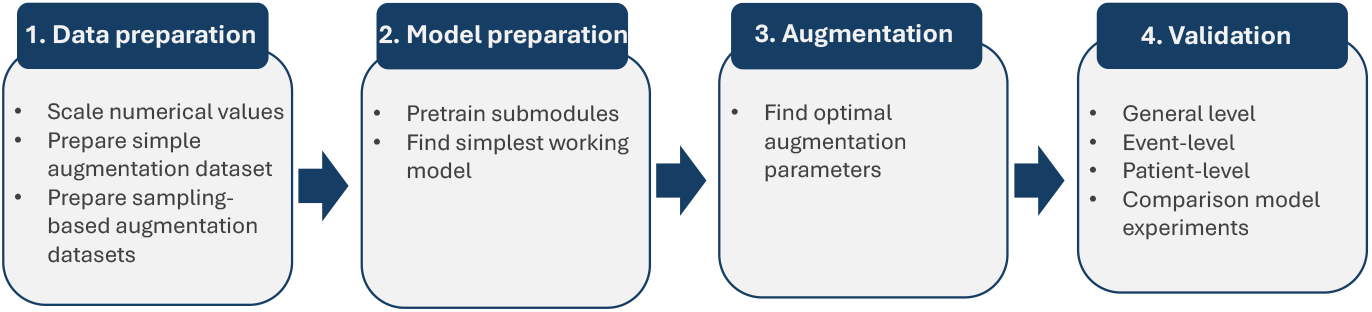
Workflow of the study.

### 2.1 Data

The dataset comprised longitudinal EHRs from patients diagnosed with any cancer at HUS Helsinki University Hospital between January 1, 2010, and December 31, 2019, with data availability extending until the end of 2024. Individuals were followed until death or five years after diagnosis, whichever occurred first.

Data included body mass indices (BMI), diagnoses, laboratory tests, medications, and procedures present within the five-year follow-up period. In addition, five static features were recorded for each patient: age at cancer diagnosis, gender, cancer diagnosis, cancer stratification [15] (Supplementary Table 2), and binary mortality label at exactly five years of follow-up.

#### 2.1.1 Preprocessing of data

Incident cancers were identified using the first three digits of the ICD-10 codes, defined by the earliest recorded cancer diagnosis (Supplementary Figures 1 and 2). Patients were excluded if their incident cancer was malignant neoplasms of independent multiple sites (C97), or if they had multiple distinct cancer diagnoses documented on the index date. To ensure diagnostic specificity, patients initially diagnosed with codes C76–C80 (malignant neoplasms of ill-defined, other secondary, and unspecified sites) were re-evaluated: if a more specific cancer code (C00–C75, C81–C96) appeared within the subsequent two months, the cancer diagnosis was updated while retaining the original date. Patients with only C76–C80 codes during this period were excluded.

Each data modality was preprocessed separately. Only events with valid timestamps were included. Diagnoses, laboratory tests, medications, and procedures present for 100 or more patients were included. Each unique diagnosis was included exactly once per patient, with the timestamp corresponding to the earliest occurrence. The other data modalities were retained longitudinally, capturing all recorded occurrences for a given patient.

The 50 most frequent numerical laboratory tests were included in the dataset. The numerical outliers were identified separately for each random variable. The mean and standard deviation (SD) were calculated across all measurements for all patients and timepoints. Outliers beyond ±3 SD were excluded per random variable. Physical units of laboratory tests were standardized. If a laboratory test had non-convertible units, it was split into two tests.

Medications were identified based on the presence of either an ATC code or a documented active ingredient of the drug. Active ingredients were subsequently mapped to their corresponding ATC codes utilizing the official Finnish Code Server registry [16].

For method development purposes, the included procedure codes were limited to surgical operations [22]. For consistency, only procedure codes with an exact length of five characters were included.

The real-valued random variables BMI and all laboratory measurements were normalized by min-max scaling across all patients and samples in the training set. The event times were measured from the first cancer diagnosis and decomposed into years, months, days, and hours.

Each event in the event sequence consisted of four parts, category, name, value, and time, respectively (Figure 2a). Event categories corresponded to the data modalities, including BMI, diagnoses, laboratory tests, medications, and procedures. Event names represented the specific medical events within a category, such as a particular diagnosis (e.g. Essential hypertension), laboratory test (e.g. hemoglobin), medication (e.g. N02AA05), and procedure (DAA00). Event sequences consisted of static information, a sequence of observed events, and padding (Supplementary Figure 6a). Patients who had 10-1000 events within a five-year follow-up period were included. The final dataset was divided into training (70%), validation (15%), and test (15%) sets using stratification of cancer diagnoses and death.

**Figure 2.**
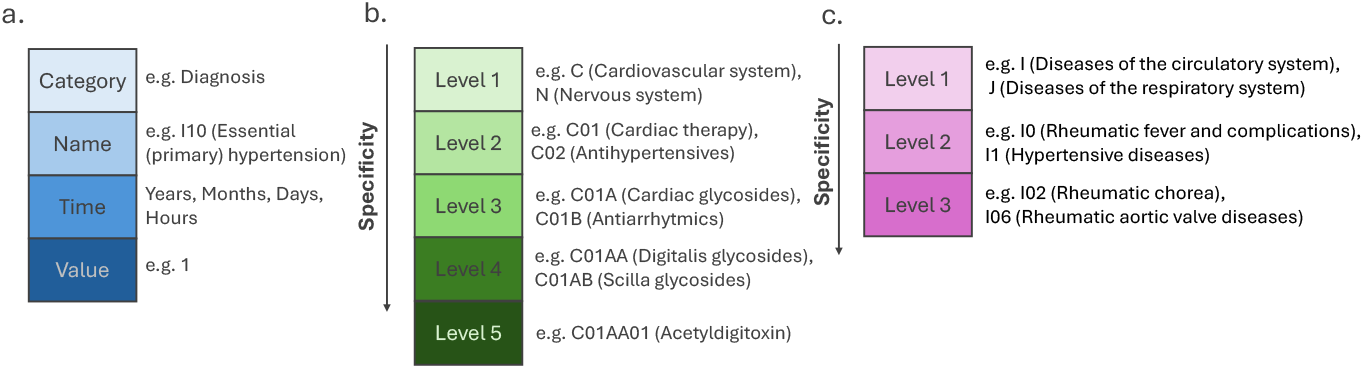
(a) Composition of each event in the event sequence. (b) Hierarchy levels of medications, ATC coding. (c) Hierarchy levels of diagnoses, ICD-10 coding.

#### 2.1.2 Data augmentation

The data augmentation protocol (Figure 3a) is applied independently to each sequence. The protocol is performed in two stages (Figure 3b).

**Figure 3.**
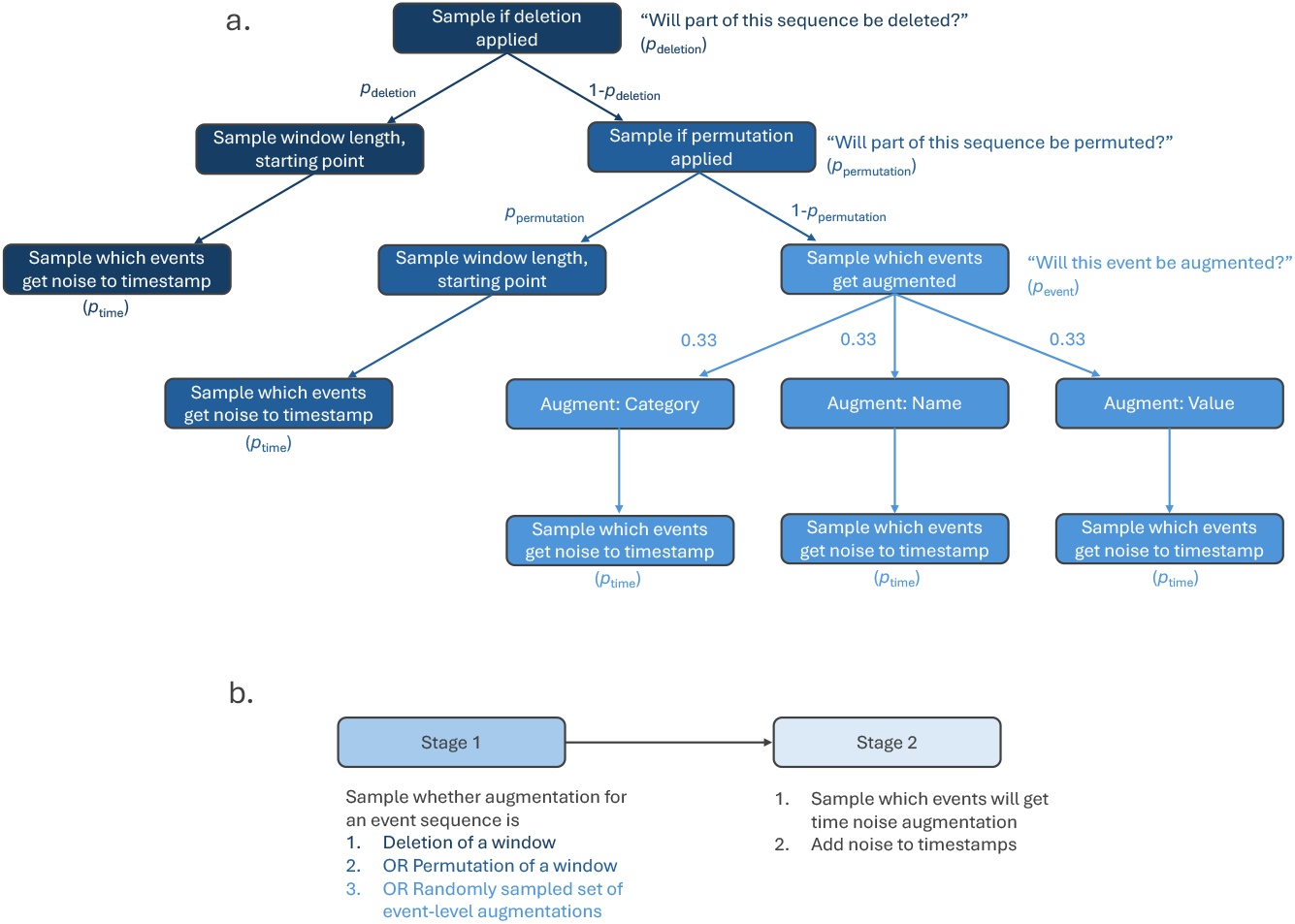
(a) The augmentation protocol as a sampling tree. The window length *w* is sampled randomly from [0.2, 0.3], corresponding to the proportion of observed events. The starting point is randomly sampled from interval [0, *M*_*i*_ − *wM*_*i*_], where *i* ∈ {1, …, *N* } indexes patients. Magnitude of noise added to timestamps is sampled from [0, max_*w*_], where max_*w*_ denotes the maximum number of weeks added to the timestamp. Refer to text and Table 1 for notations. (b) The two stages in the augmentation protocol.

In the first stage, each event sequence undergoes one of the following augmentations:

1. Deletion of part of the sequence (Supplementary Figure 4b).
2. Permutation of event order within a subsequence (Supplementary Figure 4c).
3. Event-level augmentations applied to randomly chosen events.

Here, the window lengths of deletion and permutation windows were sampled from [0.2, 0.3] to keep the augmentations moderate.

For event-level augmentations, each selected event undergoes either category augmentation, name augmentation, or value augmentation. Event name augmentations are applied such that they preserve the clinical hierarchical taxonomy of diagnosis and medication codes. (Supplementary file section 2, Supplementary Figures 3 and 4d)

In the second stage, temporal noise is added to the timestamps of a subset of events, regardless of the augmentation in the first stage (Supplementary Figure 5). The magnitude of the noise is sampled from a uniform distribution.

In the protocol, each augmentation decision is made probabilistically by sampling a value from a uniform distribution and comparing it against augmentation thresholds: probability of event sequence having a window deleted (*p*_deletion_), probability of event sequence having a window permuted (*p*_permutation_), probability of individual event being augmented (*p*_event_), and probability of individual event getting noise added to their timestamp (*p*_time_).

To identify the optimal augmentation parameters, six parameter combinations were evaluated in each stage, resulting in twelve combinations in total (Supplementary Table 3). Each combination was trained three times with different random initializations of the model, resulting in three distinct augmented datasets per combination. The final augmentation combination was selected based on the average performance across the three corresponding runs.

### 2.2 Model description

TACT builds on ClinicalTAAT [8] (Figure 4a), a transformer based model designed for irregularly sampled EHR data. It addresses irregular event timings by modeling them explicitly: Temporal information is divided into hierarchical levels (years, months, days, hours), which are embedded separately. The embedding of temporal information is independent of the embedding of other event information (category, name, value). These two channels, temporal and semantic, are integrated in Multi-head Time-aware attention (MTAA) blocks.

TACT has two main differences to ClinicalTAAT: (1) hierarchical embedding modules were added for diagnosis and medication codes, and (2) Contrastive learning framework was added following SimCLR [23] implementation with InfoNCE loss function (ℒ_InfoNCE_) [24].

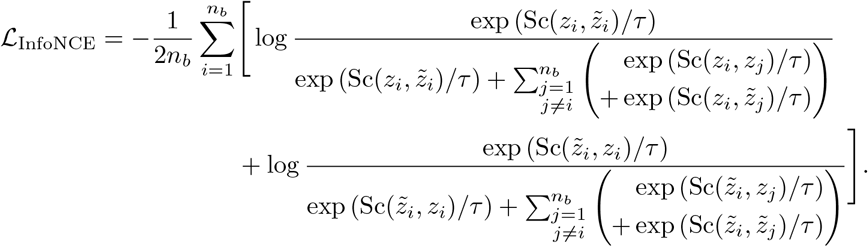

**Figure 4.**
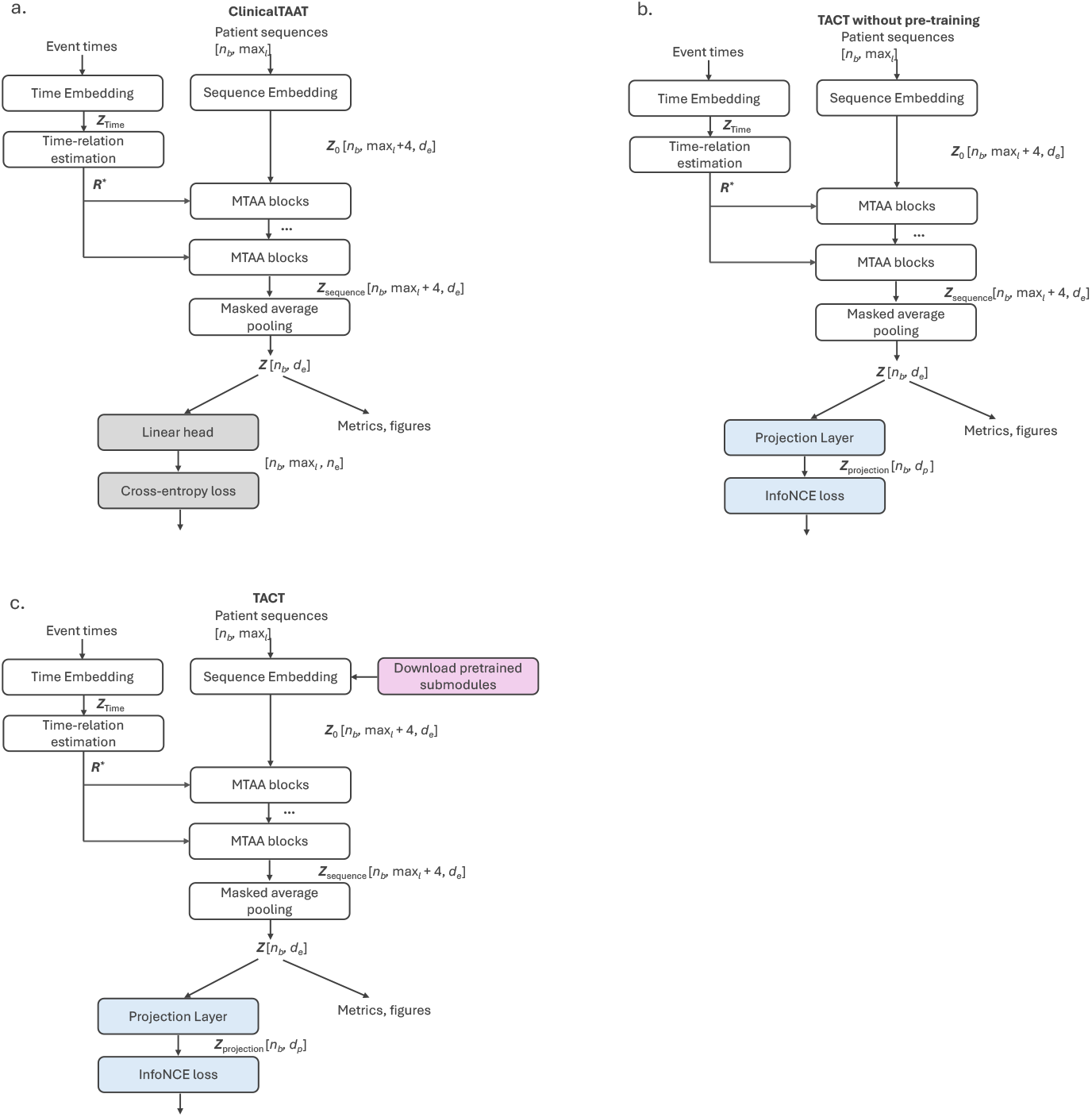
The model architectures. 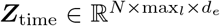 denotes the time embeddings, 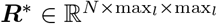 the time-relation matrices, 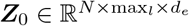 the event sequence embeddings before MTAA layers, and 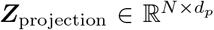 the projected patient representations. Masked-learning in grey, contrastive learning parts in blue, and pre-training of hierarchical embeddings of diagnoses and medications in pink. Refer to Table 1 for notations. (a) ClinicalTAAT. (b) TACT without pre-training. (c) TACT.

where *i, j* ∈ {1, …, *n*_*b*_ } index patients in the batch and *τ* denotes a scaling parameter. Refer to text and Table 1 for notations.

InfoNCE treats the original-augmented pair 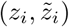 as a positive pair, and all other non-pairs as negatives. It pulls the representations of positive pairs closer in latent space while pushing the representations of non-pairs apart. The contrastive objective is symmetric: for patient *i*, (*z*_*i*_) is compared against 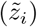 and 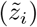 is compared against (*z*_*i*_). Due to the nature of data augmentation, the loss function inherently guides the model to produce similar representations for similar event sequences and dissimilar representations for dissimilar ones. Following the SimCLR framework [23], a non-linear projection layer is applied to patient representation 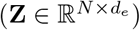 before computing the loss (ℒ_InfoNCE_).

TACT embeds each semantic component of an event (category, name, and value) separately (Supplementary Figure 6b). The event values are embedded using continuous value embeddings. The event categories are embedded using a lookup table, where each category is first mapped to an index and then represented as real-valued vectors using a PyTorch `nn.Embedding` layer. The event names for BMI, laboratory tests, and procedures are embedded in a similar way. Diagnoses and medications are embedded using separate submodules (Supplementary Figures 7 and 8). Each level of the diagnosis and medication code hierarchy (Figure 2b-c) is embedded separately, and finally summed, preserving the clinical taxonomy of ICD-10 and ATC codes. The submodules are pre-trained prior to TACT training and fine-tuned during training. To achieve the embedding of each observed event, the separately-embedded components are summed: 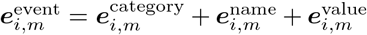. Here, *m* ∈ {1, …, *M*_*i*_} indexes the events of patient *i*. The embedding vector 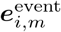 denotes the embedding of *m*-th event of patient *i*. Refer to Table 1 for notations.

Patient representations **Z** are obtained from contextualized event sequence representations **Z**_sequence_ via masked average pooling over observed events. All metrics and figures in this study are produced using **Z**. Refer to text and Table 1 for notations.

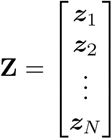

### 2.3 Comparison models

ClinicalTAAT was used as baseline comparison model. The second baseline comparison model was TACT without pre-training (Figure 4b), which lacks the hierarchical pre-training submodules.

### 2.4 Implementation details

For TACT and the two comparison models the following implementations - differing from original ClinicalTAAT implementation - were applied: (1) The temporal granularity was extended to include years, months, days, and hours. (2) The feed-forward network (FFN) in the attention blocks was replaced with a GeGLU layer [25], a variant of a gated linear unit (GLU). (3) The event embeddings consisted of three components (category, name, value). (4) The static features were added as the four first events for each event sequence. (5) All bias terms were set to False.

Each model was trained for up to 300 epochs with early stopping (patience=20, delta=10^−4^), and the same hyperparameter configuration (Supplementary Figure 9) was used across all models.

### 2.5 Evaluation metrics

The model performance was evaluated at two levels: event embeddings and patient representations. Event embedding quality was assessed based on (i) coherence of event embeddings within each event category and (ii) distinctiveness between categories. Patient representation quality was measured as the similarity within original-augmented pair and dissimilarity between non-pairs. Here, in event level analysis classes correspond to the event categories and samples individual events. In patient level, each class refers to an original-augmented pair.

Distinctiveness of representations (DOR) measures the embedding quality with the average cosine similarity within (DOR_within_class_, DOR_within_patient_) and between classes (DOR_across_class_, DOR_across_patient_). DOR can be applied for event categories and augmentation pairs.

For the class level, DOR_within_class_ and DOR_across_class_:

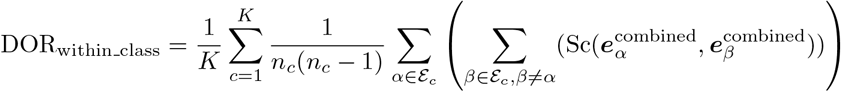

where *c* ∈ {1, …, *K*} indexes the classes. The indices *α, β* ∈ {1, …, *n*_*c*_} index event embeddings in class *c*. 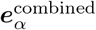 and 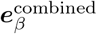 denote the corresponding combined event embeddings. Refer to text and Table 1 for notations.

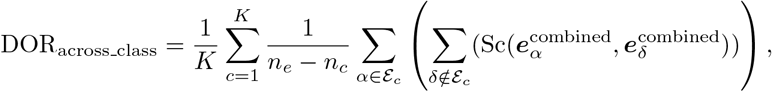

where the index *δ* ∈ {1, …, *n*_*e*_ − *n*_*c*_ } indexes samples outside class *c*, and 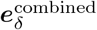 denotes corresponding combined event embeddings.

For the patient level, DOR_within_patient_ and DOR_across_patient_:

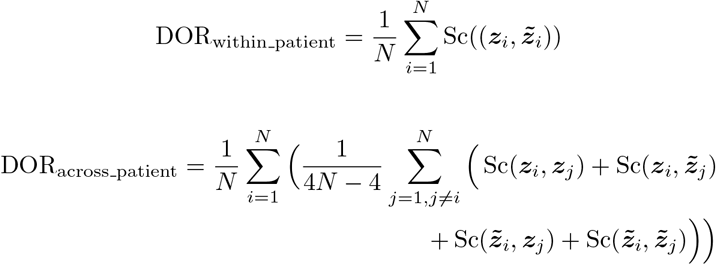

As an extension of DOR, accuracy of distinctiveness of representations (ADOR_class_, ADOR_patient_) quantifies the class separation in the embedding space. The metric is inspired by sample alignment accuracy (SAA), which measures euclidean distances instead of cosine similarities [26].

For the class level, ADOR_class_:

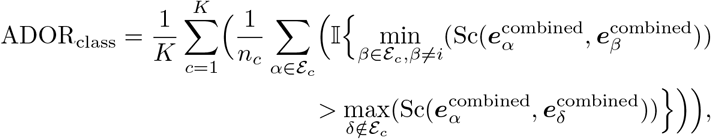

where I(*·*) denotes the indicator function.

For the patient level, ADOR_patient_:

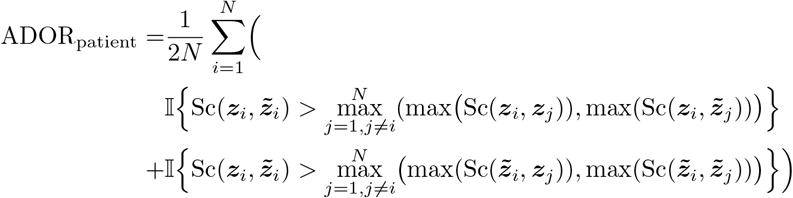

ADOR is the proportion of samples whose embeddings are more similar to those within the same class than to those from different classes. Compared to DOR, ADOR (range [0,1]) provides an intuitive interpretation of the embedding space. At the patient level, ADOR is the proportion of pairs whose original and augmented representations are more similar to each other than to other patients’ representations and their respective augmentations.

Class alignment accuracy (CAA) describes class separation in the embedding space. It is the proportion of samples with a smaller euclidean distance within the class compared to samples from other classes. [26]

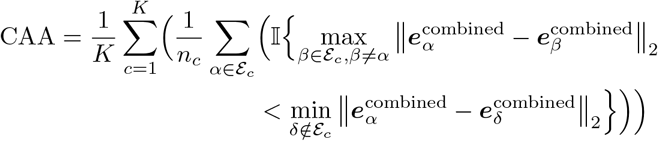

Class alignment consistency (CAC) measures the coherence of neighborhoods in the embedding space and represents the proportion of samples for which their *k* nearest neighbors belong to the same class as the sample itself. [26].

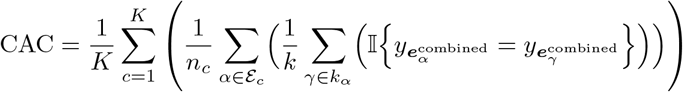

where *k* denotes the number of neighbors, *k*_*α*_ the set of *k* nearest neighbors of the combined event embedding 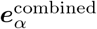, and 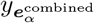 denotes the category of this embedding. The index *γ* ∈ *k*_*α*_ indexes neighboring combined event embeddings, and 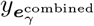 denotes the category of the corresponding combined event embedding.

Here, *k* is computed as the proportion (%) of all available samples. To ensure a fair comparison of neighborhood similarities of classes, we set *k* to min(*k*_class_, *k*). *k*_class_ is the number of samples in the corresponding class.

## 3 Results

### 3.1 Dataset characteristics

The final dataset comprised 77,628 patients, of whom 53% were females. The median follow-up time was 60 months for all patients and 12 months among patients who died (29%) (Table 2, Supplementary Table 5, Supplementary Figures 10 and 11)

**Table 2.** Cohort characteristics. Values presented as medians or n (%) unless otherwise specified. Interquantile Range (IQR) is the difference between quartile 1 (Q1) and quartile 3 (Q3).

| Characteristic | Value | IQR [Q1, Q3] | Unit |
| --- | --- | --- | --- |
| Patients | 77 628 | - | n |
| Female | 41 418 (53%) | - | n |
| Age at diagnosis | 68 | 18 [58, 76] | Years |
| Cancer diagnoses | 82 | - | n |
| Sequence length | 188 | 326 [75, 401] | n |
| Death within 5 years | 22 781 (29%) | - | n |
| Follow-up time | 60 | 21 [39, 60] | Months |
| Time to death | 12 | 24 [4, 28] | Months |
| Time to last event (or death) | 47 | 40 [17, 58] | Months |

### 3.2 Augmentation experiments

TACT maintained stable performance across the twelve augmentation combinations (Supplementary Table 3) in validation loss and patient-level evaluation metrics. Notable performance variations were observed in event-level metrics, which were subsequently used to select the optimal hyperparameter baseline (Supplementary Tables 6 and 7).

An augmentation dataset with extreme temporal and semantic noise was used to evaluate the robustness of TACT. TACT maintained event- and patient-level performance comparable to results achieved with moderate augmentation parameters (Supplemantary Table 8 and Supplementary Figure 12, Augmentation combination 12, Supplementary Table 7).

### 3.3 Event-Level Analysis

TACT learned distinct and well-separated embeddings for each event category (Figure 5a-b, Table 4). Crucially, the inherent clinical hierarchies of ICD-10 (diagnosis) and ATC (medication) codes were accurately preserved. Codes sharing the same initial character exhibited substantially higher cosine similarity with each other than with codes from unrelated categories, resulting in clear and coherent clustering across diagnostic and medication groups (Figure 5c-e). For medication codes, the hierarchical taxonomy was maintained consistently down to the sub-levels (Figure 5f).

**Table 3.** Number of recorded observations by data modality.

| Data modality | Unique variables | Patients with data | Total count |
| --- | --- | --- | --- |
| BMI | 1 | 56 637 (73%) | 303 435 |
| Diagnoses | 601 | 67 851 (87%) | 357 370 |
| Laboratory tests | 50 | 69 112 (89%) | 14 950 465 |
| Medications | 733 | 74 045 (95%) | 5 173 857 |
| Procedures | 308 | 54 555 (70%) | 204 989 |

**Table 4.**
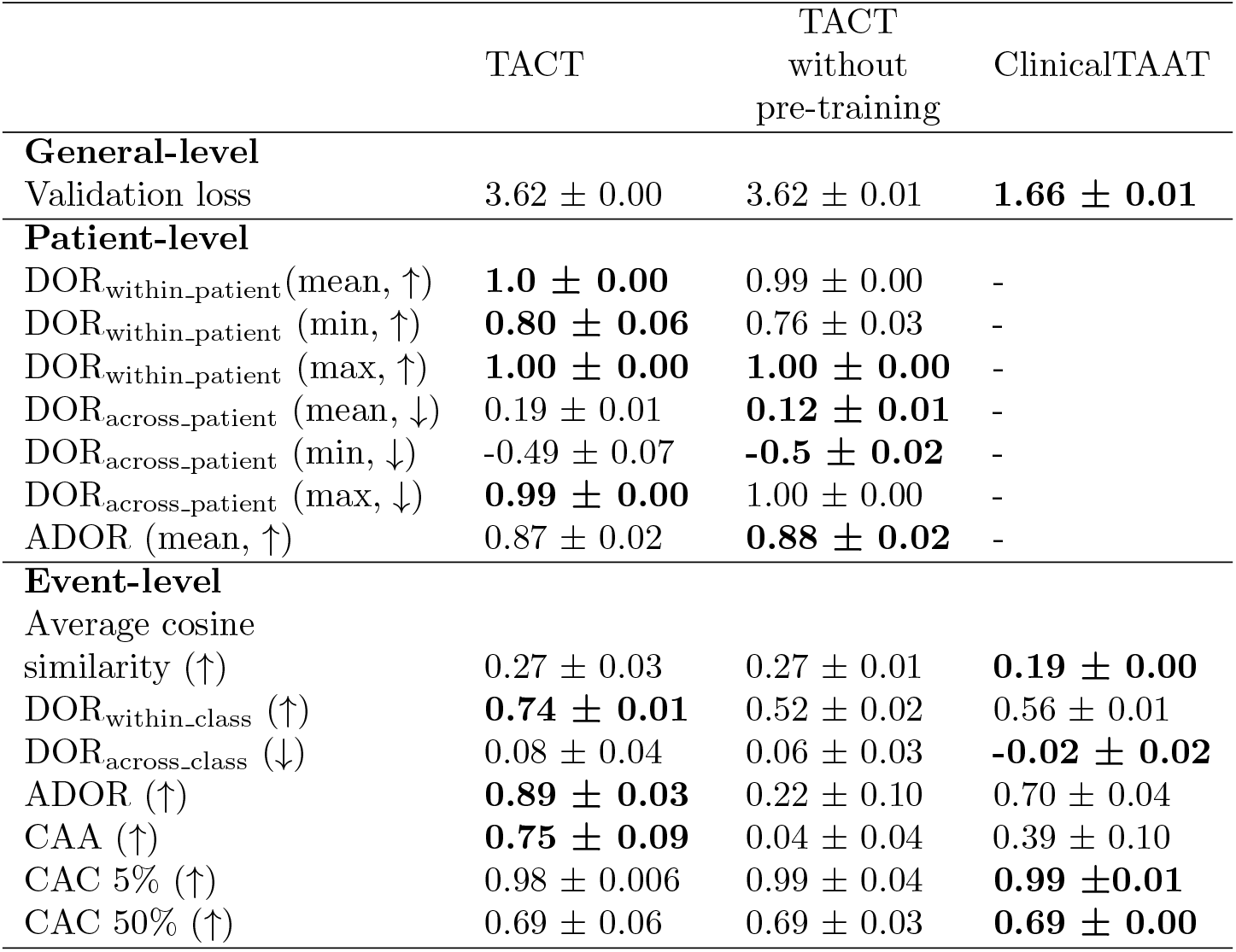
The metrics on test data. The best performance is bolded. Values reported as mean ± SD. ↑ denotes that higher value is better, ↓ that low value is better. “mean”, “min”, and “max” here refer to the average, minimum, and maximum value recorded for the corresponding metric. CAC % denotes the proportion of all available samples included in the neighborhood.

**Figure 5.**
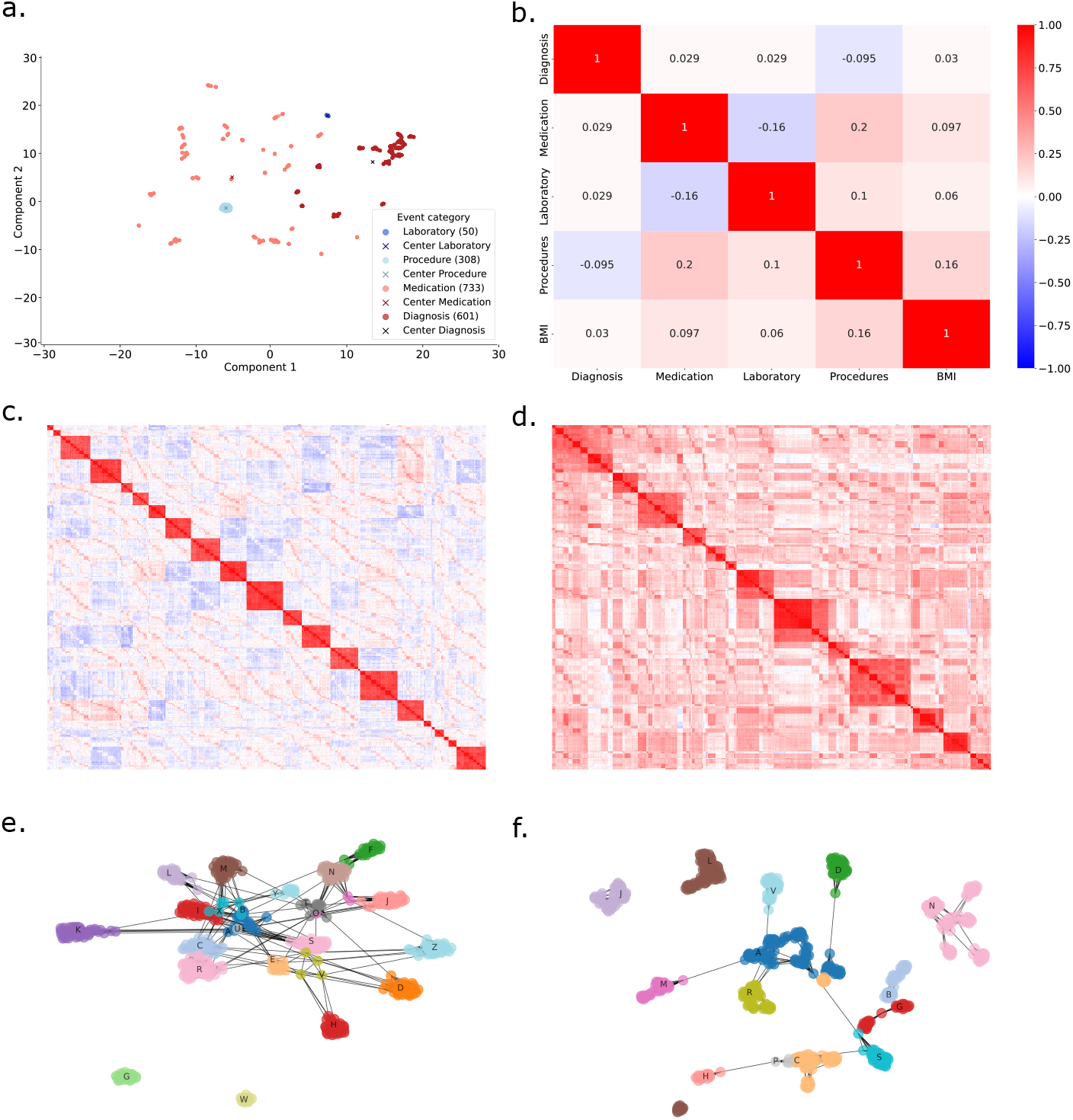
Results of event-level performance of TACT. (a) The combined event embeddings ***e***^combined^ projected into 2D with UMAP. Refer to Table 1 for notations. (b) The cosine similarities of event category embeddings. (c) Cosine similarity heatmap of diagnosis embeddings. (d) Cosine similarity heatmap of medication embeddings. (e) Hierarchical clustering of learned diagnosis embeddings. (f) Hierarchical clustering of learned medication embeddings.

### 3.4 Patient-Level Analysis

TACT learned highly similar representations for original-augmented pairs, while the representations of non-pairs remained moderately similar (Figure 6a). Latent space analysis revealed a well-structured topological organization of patient representations, effectively mapping similar patient representations close to one another (Figure 6b).

**Figure 6.**
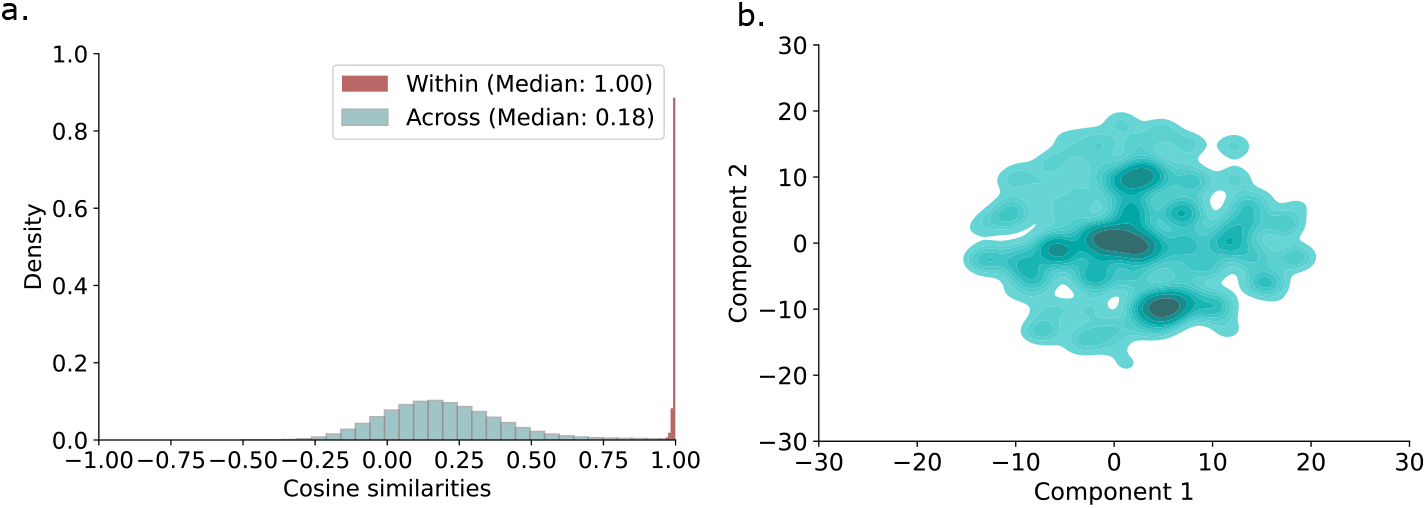
The patient-level results of TACT on test data. (a) Cosine similarities of patient representations and augmentation representations. Both within original-augmented similarities and across similarities (non-pairs). (b) The latent space of patient representations, projected into 2D with UMAP.

### 3.5 Model comparison

To isolate the specific contributions of individual model components, TACT was compared with baseline comparison models, ClinicalTAAT and TACT without pre-training.

The absence of the contrastive learning framework in ClinicalTAAT caused the patient representation space to collapse into a single dense region (Figure 7e). In contrast, ClinicalTAAT learned more distinct combined event embeddings ***e***^combined^ than the other two models, resulting in improved class separation (Figures 5b and 7a-b, average cosine similarity and DOR_across_class_ in Table 4) but reduced coherence within categories (DOR_within_class_, ADOR, and CAA in Table 4). The increase in neighborhood similarity (CAC in Table 4) was not significant.

**Figure 7.**
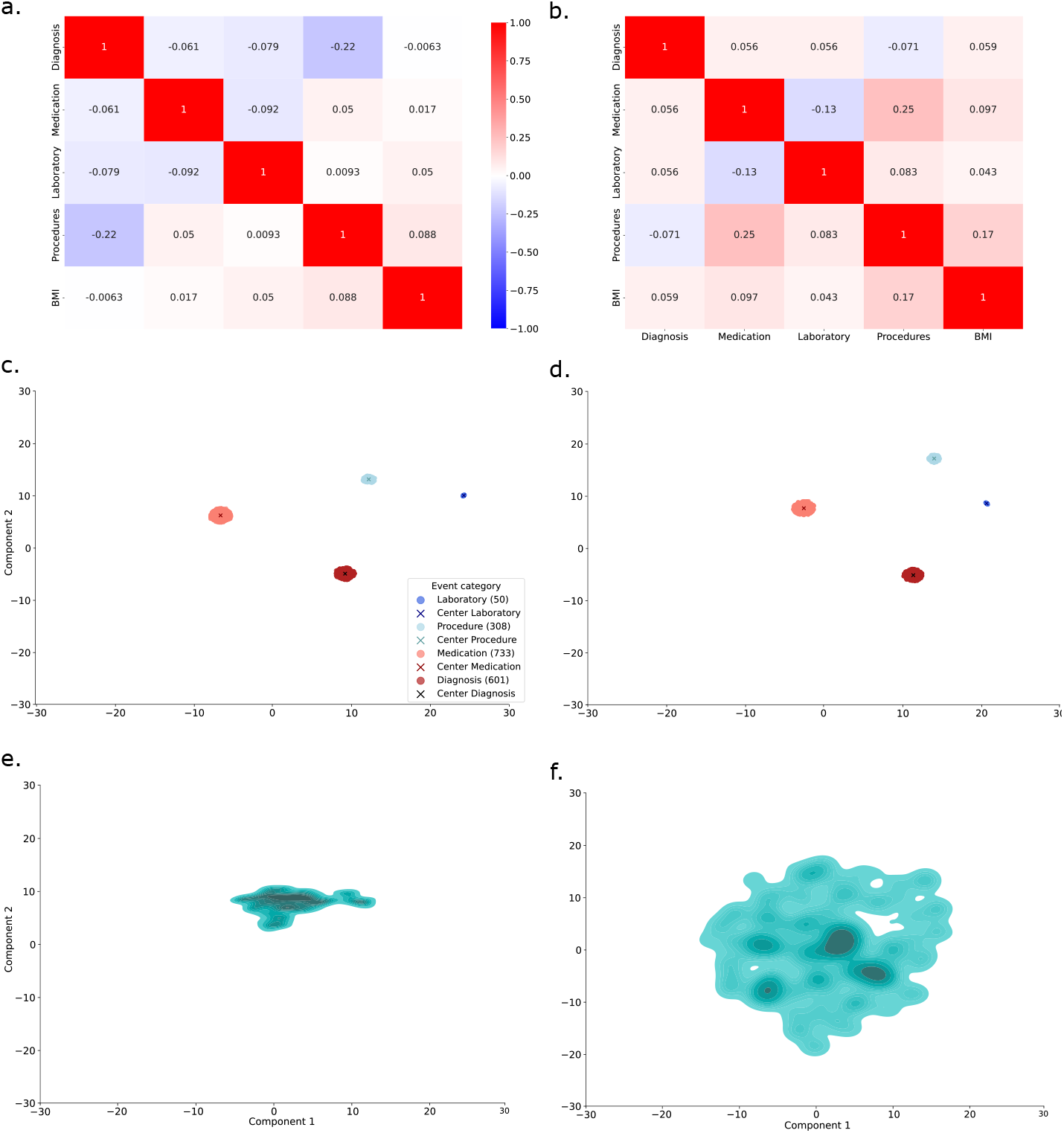
The comparison model results. Refer to Table 1 for notations. (a) The cosine similarities of event category embeddings, ClinicalTAAT. (b) The cosine similarities of event category embeddings, TACT without pre-training. (c) The combined event embeddings ***e***^combined^ projected into 2D with UMAP, ClinicalTAAT. (d) The combined event embeddings ***e***^combined^ projected into 2D with UMAP, TACT without pre-training. (e) The latent space of patient representations of test data, projected into 2D with UMAP, ClinicalTAAT. (f) The latent space of patient representations of test data, projected into 2D with UMAP, TACT without pre-training.

The absence of hierarchical pre-training resulted in a complete collapse of the hierarchical taxonomy in the event embedding space: unlike TACT, the comparison models failed to map the ICD-10 and ATC codes into their clinically relevant biological groupings (Figure 7c-d). Moreover, event-level metrics were significantly lower for TACT without pre-training than with the complete architecture (Event-level metrics in Table 4). This effect was further supported by categorical breakdown: event categories subjected to hierarchical pre-training, i.e., diagnoses and medications, exhibited substantially higher metrics compared to laboratory tests and procedures (Supplementary Table 9). In contrast, the absence of a pre-training component (TACT without pre-training) did not significantly affect the structure of the latent space of patient representations (Figure 7f). A slight decrease in non-pair cosine similarity was observed, as well as a slight decrease in within pair similarity (DOR_across_patient_ (min, mean) and DOR_within_patient_ (min, mean) in Table 4).

## 4 Discussion

The main objective of this study was to develop a framework for learning high-quality patient representation vectors without relying on external labels for downstream tasks. The proposed framework leverages contrastive learning to learn accurate patient representations, captures temporal and semantic relationships through a time-aware transformer architecture, and addresses data irregularity and sparsity using hierarchical time embeddings and explicit time relation modeling.

Our augmentation protocol provides a versatile approach for augmenting event sequences from structured EHR data. It combines existing techniques, such as order permutation [12],[14] and value noise injection [13], with new components. These include deletion of event windows and the use of clinical hierarchies of diagnosis and medication codes for event replacement. The sampling-based design further enhances flexibility and robustness. Unlike previous approaches, which either apply identical augmentations to all sequences [14],[13] or limit augmentation to a single type per event sequence [12], our protocol enables multiple augmentation types per event sequence and performs augmentations independently for each event sequences. Additionally, the proposed category- and hierarchy-based event replacement is more straightforward than current approaches that use cosine similarity-based synonym insertion [12].

The experiments demonstrated that TACT effectively compresses irregular and longitudinal EHRs event sequences into structured patient representations. The learned patient representations reflected the characteristics of the dataset: cosine similarities of the original-augmented pairs ranged from 0.80 to 1, while similarities between non-pairs were centered around 0.2. The distribution of the non-pair cosine similarities indicates that the majority of the learned representations were only moderately similar, with a small fraction of non-pairs being highly similar. This pattern is likely driven by the predominance of laboratory tests in the dataset. Patients with recurring laboratory patterns are likely to increase the non-pair cosine similarities due to the similar clinical patterns. This observation is further supported by the topology of the latent space. Multiple highly dense regions are separated by sparser intervals, which captures the complexity and continuity of patient trajectories. The robustness and stability of TACT were further demonstrated using extremely noisy input data.

The model comparison experiments demonstrated the complementary roles of hierarchical pre-training and contrastive learning. Hierarchical embedding improved event-level performance and embedding quality, while contrastive learning enhanced the patient representation learning. These differences are explained by the distinct training objectives: ClinicalTAAT emphasizes learning contextual relationships in data, whereas contrastive learning focuses on learning similar representations for similar event sequences. This distinction is evident in the latent space: TACT and TACT without pre-training produce a structured latent space, while the patient representations of ClinicalTAAT collapse into a single dense region. In summary, contrastive learning improves patient representation quality, but requires hierarchical pre-training to achieve comparable event-level performance. Together, these observations highlight the importance of the complete TACT architecture.

The limitations of this study include the use of a single dataset and the lack of extensive clustering analysis to characterize the patient representation subgroups identified by TACT. Additionally, the relatively small batch size used during training may have limited the models’ ability to learn more expressive representations.

Future work should explore augmentation of static features (e.g. age, gender, cancer diagnosis, cancer stratification) and the incorporation of the hierarchy of procedure codes. The effects of per-epoch augmentation and sequence length should also be evaluated. Finally, the framework should be applied to identify patient subgroups based on disease progression.

In conclusion, we introduced an augmentation workflow tailored for learning generalizable, longitudinal patient representations from multimodal EHR data, leveraging the hierarchical structure of diagnosis and medication codes. This sampling-based protocol provides an efficient and flexible approach for generat-ing diverse augmentations. We demonstrated that TACT learns high-quality, structured event embeddings and patient representations. The combination of hierarchical pre-training and contrastive learning outperforms masked learning, ensuring both high-quality event embeddings and informative patient representations.

## Data Availability

The data is not publicly available due to national legislation of individual level clinical data, which was used with the permission of HUS Helsinki University Hospital. For data permission inquiries, please contact.

## 5 Contributions

A.H. prepared clinical data for analysis, performed analyses and was the lead author. A.H and L.T performed methodology development and visualization. A.H., L.T. and A.A contributed to software development. M.K. and R.R. launched and supervised the study. L.T. contributed to writing. A.H., L.T., A.A., S.S., R.R. and M.K. contributed to the revision of the manuscript.

## 6 Code Availability

The implementation of the proposed framework and comparison models are available at the public repository [upon publishing] under MIT license.

