## Supplementary_file for "Time-Aware Contrastive Transformer for Longitudinal Patient Representation Learning"

Anni Heinolainen (0009-0007-0690-8844) [1,2,*],
Linh Tran (0009-0007-2846-1342) [1],
Arushi Arora (0009-0005-0234-1372) [1,2],
Suvi Silén (0000-0003-1240-0661)[1,2],
Risto Renkonen (0000-0003-4455-2918)[1,2],
Miika Koskinen (0000-0002-7267-5811)[1,2,3]

[1] Faculty of Medicine, University of Helsinki, Helsinki, Finland; [2] HUS Diagnostic Center, Helsinki University Hospital, Helsinki, Finland; [3] New Children's Hospital, Helsinki University Hospital, Helsinki, Finland.

The mathematical variables used in the Supplementary file are listed in Supplementary Table 1.

Supplementary Table 1: Summary of mathematical variables used in the supplementary file.

| **Variable** | **Definition** |
|---|---|
| $N$ | Number of patients |
| $d_e$ | Embedding dimension |
| $d_p$ | Projection dimension |
| $e \in \mathbb{R}^{d_e}$ | Embedding of an event |
| $\uparrow$ | Denotes that higher value is better |
| $\downarrow$ | Denotes that lower value is better |

### 1 Details of the final dataset

Supplementary Table 2: Cancer stratifications and included cancer diagnoses. Stratification based on ICD-10 hierarchy [1].

| Cancer stratification | Cancers (ICD-10) |
|---|---|
| Lip, oral cavity, and pharynx | C00-C14 |
| Digestive organs | C15-C26 |
| Respiratory and intrathoracic organs | C30-C39 |
| Melanoma and skin neoplasm | C43-C44 |
| Mesothelial and soft tissue neoplasm | C45-C49 |
| Breast | C50 |
| Female genital organs | C51-C58 |
| Male genital organs | C60-C63 |
| Urinary tract | C64-C68 |
| Eye, brain and other parts of central nervous system | C69-C72 |
| Thyroid and other endocrine glands | C73-C75 |
| Neoplasms, stated of presumed to be primary of lymphoid, hematopoietic, and related tissue | C81-C96 |

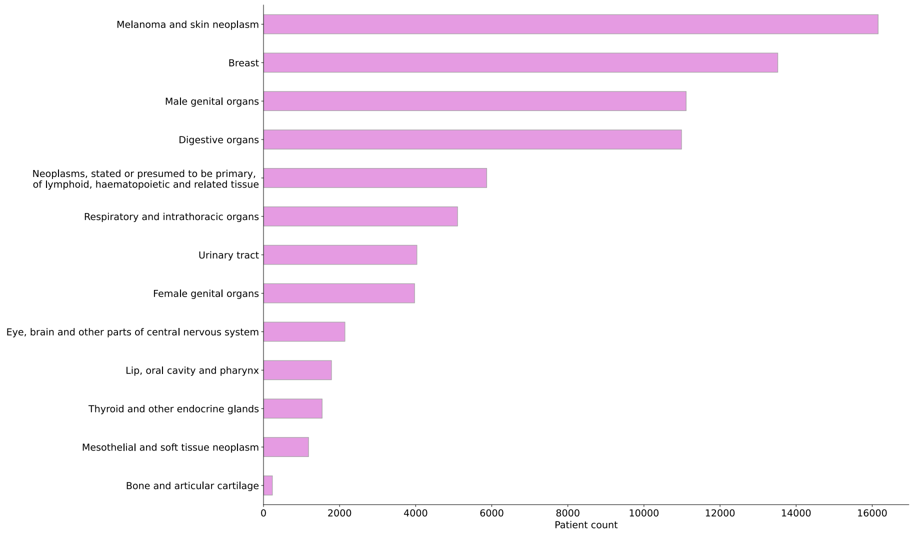


Supplementary Figure 1: The distribution of the stratifications [1] of primary cancer diagnosis.

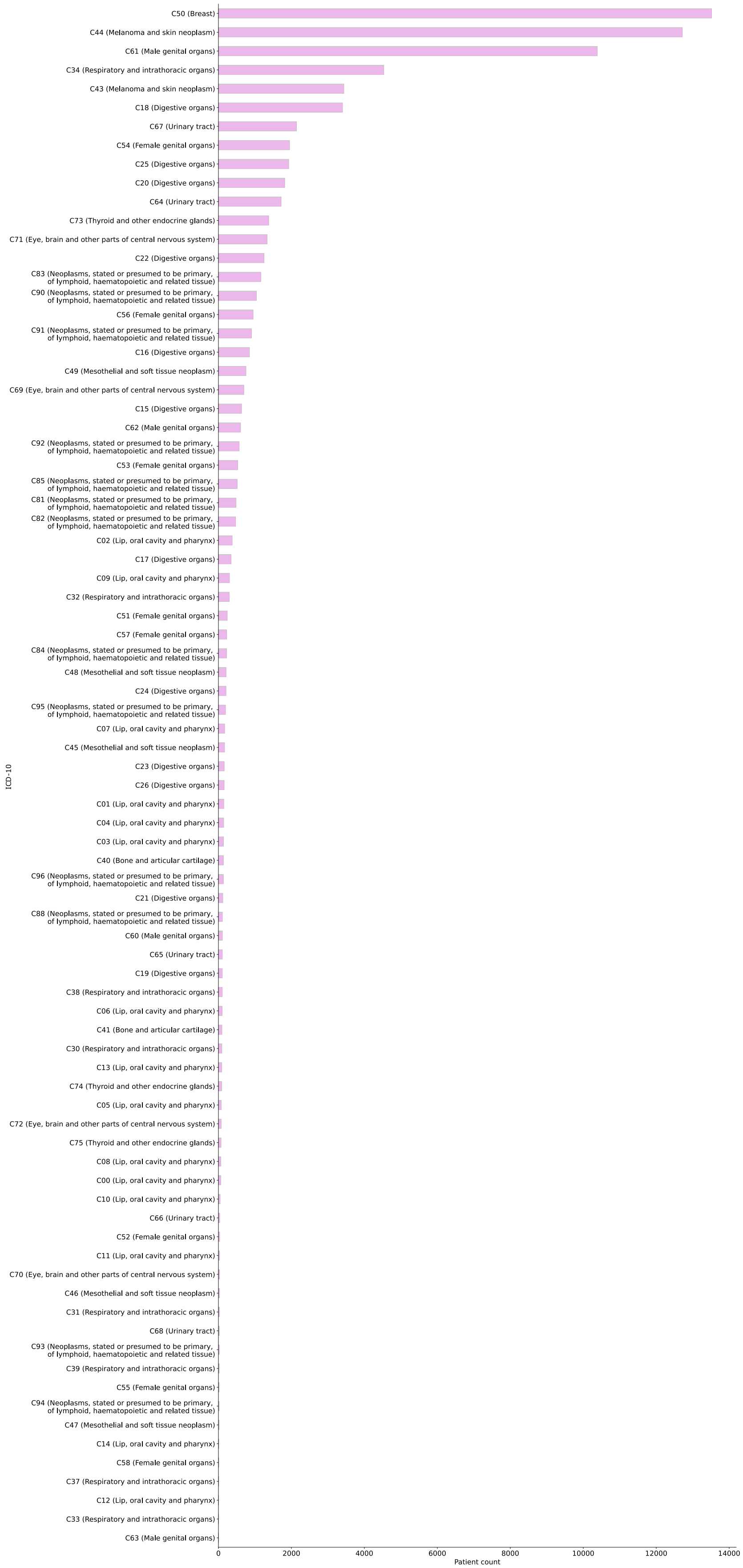

C50 (Breast)
C44 (Melanoma and skin neoplasm)
C61 (Male genital organs)
C34 (Respiratory and intrathoracic organs)
C43 (Melanoma and skin neoplasm)
C18 (Digestive organs)
C67 (Urinary tract)
C54 (Female genital organs)
C25 (Digestive organs)
C20 (Digestive organs)
C64 (Urinary tract)
C73 (Thyroid and other endocrine glands)
C71 (Eye, brain and other parts of central nervous system)
C22 (Digestive organs)
C83 (Neoplasms, stated or presumed to be primary, of lymphoid, haematopoietic and related tissue)
C90 (Neoplasms, stated or presumed to be primary, of lymphoid, haematopoietic and related tissue)
C56 (Female genital organs)
C91 (Neoplasms, stated or presumed to be primary, of lymphoid, haematopoietic and related tissue)
C16 (Digestive organs)
C49 (Mesothelial and soft tissue neoplasm)
C69 (Eye, brain and other parts of central nervous system)
C15 (Digestive organs)
C62 (Male genital organs)
C92 (Neoplasms, stated or presumed to be primary, of lymphoid, haematopoietic and related tissue)
C53 (Female genital organs)
C85 (Neoplasms, stated or presumed to be primary, of lymphoid, haematopoietic and related tissue)
C81 (Neoplasms, stated or presumed to be primary, of lymphoid, haematopoietic and related tissue)
C82 (Neoplasms, stated or presumed to be primary, of lymphoid, haematopoietic and related tissue)
C02 (Lip, oral cavity and pharynx)
C17 (Digestive organs)
C09 (Lip, oral cavity and pharynx)
C32 (Respiratory and intrathoracic organs)
C51 (Female genital organs)
C57 (Female genital organs)
C84 (Neoplasms, stated or presumed to be primary, of lymphoid, haematopoietic and related tissue)
C48 (Mesothelial and soft tissue neoplasm)
C24 (Digestive organs)
C95 (Neoplasms, stated or presumed to be primary, of lymphoid, haematopoietic and related tissue)
C07 (Lip, oral cavity and pharynx)
C45 (Mesothelial and soft tissue neoplasm)
C23 (Digestive organs)
C26 (Digestive organs)
C01 (Lip, oral cavity and pharynx)
C04 (Lip, oral cavity and pharynx)
C03 (Lip, oral cavity and pharynx)
C40 (Bone and articular cartilage)
C96 (Neoplasms, stated or presumed to be primary, of lymphoid, haematopoietic and related tissue)
C21 (Digestive organs)
C88 (Neoplasms, stated or presumed to be primary, of lymphoid, haematopoietic and related tissue)
C60 (Male genital organs)
C65 (Urinary tract)
C19 (Digestive organs)
C38 (Respiratory and intrathoracic organs)
C06 (Lip, oral cavity and pharynx)
C41 (Bone and articular cartilage)
C30 (Respiratory and intrathoracic organs)
C13 (Lip, oral cavity and pharynx)
C74 (Thyroid and other endocrine glands)
C05 (Lip, oral cavity and pharynx)
C72 (Eye, brain and other parts of central nervous system)
C75 (Thyroid and other endocrine glands)
C08 (Lip, oral cavity and pharynx)
C00 (Lip, oral cavity and pharynx)
C10 (Lip, oral cavity and pharynx)
C66 (Urinary tract)
C52 (Female genital organs)
C11 (Lip, oral cavity and pharynx)
C70 (Eye, brain and other parts of central nervous system)
C46 (Mesothelial and soft tissue neoplasm)
C31 (Respiratory and intrathoracic organs)
C68 (Urinary tract)
C93 (Neoplasms, stated or presumed to be primary, of lymphoid, haematopoietic and related tissue)
C39 (Respiratory and intrathoracic organs)
C55 (Female genital organs)
C94 (Neoplasms, stated or presumed to be primary, of lymphoid, haematopoietic and related tissue)
C47 (Mesothelial and soft tissue neoplasm)
C14 (Lip, oral cavity and pharynx)
C58 (Female genital organs)
C37 (Respiratory and intrathoracic organs)
C12 (Lip, oral cavity and pharynx)
C33 (Respiratory and intrathoracic organs)
C63 (Male genital organs)
ICD-10
0
2000
4000
6000
8000
10000
12000
14000
Patient count

#### 2 Data Augmentation details

The augmentation protocol is applied independently for each event sequence. The action in each step of the protocol is determined in sampling-based manner by drawing random numbers and comparing them against a threshold. The steps in the protocol are as follows:

1. First, for each sequence, a value from [0,1] is sampled to determine whether it undergoes a deletion. If the drawn value exceeds the deletion threshold ($p_{\text{deletion}}$), a part of the sequence is removed.

2. Otherwise, a new value is sampled. Similarly, if it exceeds a permutation threshold ($p_{\text{permutation}}$), the order of a subset of events is changed.

3. If neither of the conditions is met, event-level augmentations are applied to a subset of the events.

4. Finally, a time augmentation is performed for a subset of the events for each sequence regardless if it underwent deletion, permutation or individual event augmentations.

In both deletion and permutation augmentations, the window length (proportion of events within an event sequence), is sampled uniformly from [0.2, 0.3]. These limits can be changed, but here were chosen to keep the window-sizes moderate. The starting point of the window is randomly sampled. Deletion removes all events within the specified window (Supplementary Figure 4b), whereas permutation changes the order of the events while fixing the timestamps (Supplementary Figure 4c).

In individual event level augmentations, each event of an event sequence is assigned a randomly sampled value from [0,1]. If the value exceeds the event augmentation threshold ($p_{\text{event}}$), an augmentation type (category, name, value) is sampled uniformly. In event category augmentation, event category, name, and event value are modified (Supplementary Figure 3a). In event name augmentation, event name and event value are modified (Supplementary Figure 3 b). In event value augmentation, only the event value is modified (Supplementary Figure 3d).

In all augmentation types, the event values are set to 1 for diagnoses, medications, and procedures. For BMI and laboratory tests, the new event values are randomly sampled from [0,1] in category and name augmentations. In event value augmentations, noise from a Gaussian distribution ($\mathcal{N} \sim (0, 0.05)$) is added to the event values of BMI and laboratory measurements (Supplementary Figure 3e).

The event names are augmented by sampling from the same category (Supplementary Figure 3c). For laboratory measurements and procedures, a new event name is sampled uniformly until it is not the original one. For diagnosis and medications, hierarchical coding systems (ICD-10 and ATC) are leveraged. The augmentation is performed in two steps: (1) augmentation level is sampled from a categorical distribution (Level 1 - Level 3 for diagnoses, Level 1 - Level 4 for medications, Supplementary Table 4). (2) Candidate sets for event name sampling are isolated based on the sampled hierarchy level. The original event name is deleted from the candidate sets if possible. The candidate sets are constructed as follows:

1. Level 1: The new code begins with a different character than the original code.
2. Level 2: Diagnoses share the first character but differ on the second. Medications share the first character but differ within the first triplet. If empty, the first two (for diagnoses) or three (for medications) characters may match.
3. Level 3: Diagnoses share the first two characters. Medications share the first triplet but differ on the fourth character. If empty, the first four characters may match.
4. Level 4 (medications only): The first four characters remain identical.

In category-level augmentation, a new event category is first randomly sampled from the remaining categories. Second, a new event name is uniformly sampled from the corresponding set of event names (Supplementary Figure 3b).

In time noise augmentation, for each event in all sequences, a value from [0,1] is sampled. If it exceeds a threshold ($p_{\text{time}}$), noise is added to the individual event. The amount of noise added to the timestamps is sampled from $[0, \max_w]$, where $\max_w$ denotes the maximum number of weeks added to a timestamp.

Here, the augmentation parameters were optimized in two steps. First, $p_{\text{deletion}}$, $p_{\text{permutation}}$, and $p_{\text{event}}$ were evaluated. Second, $p_{\text{time}}$ and $\max_w$ were tuned (Supplementary Table 4). Both steps used six parameter combinations (Supplementary Table 3).

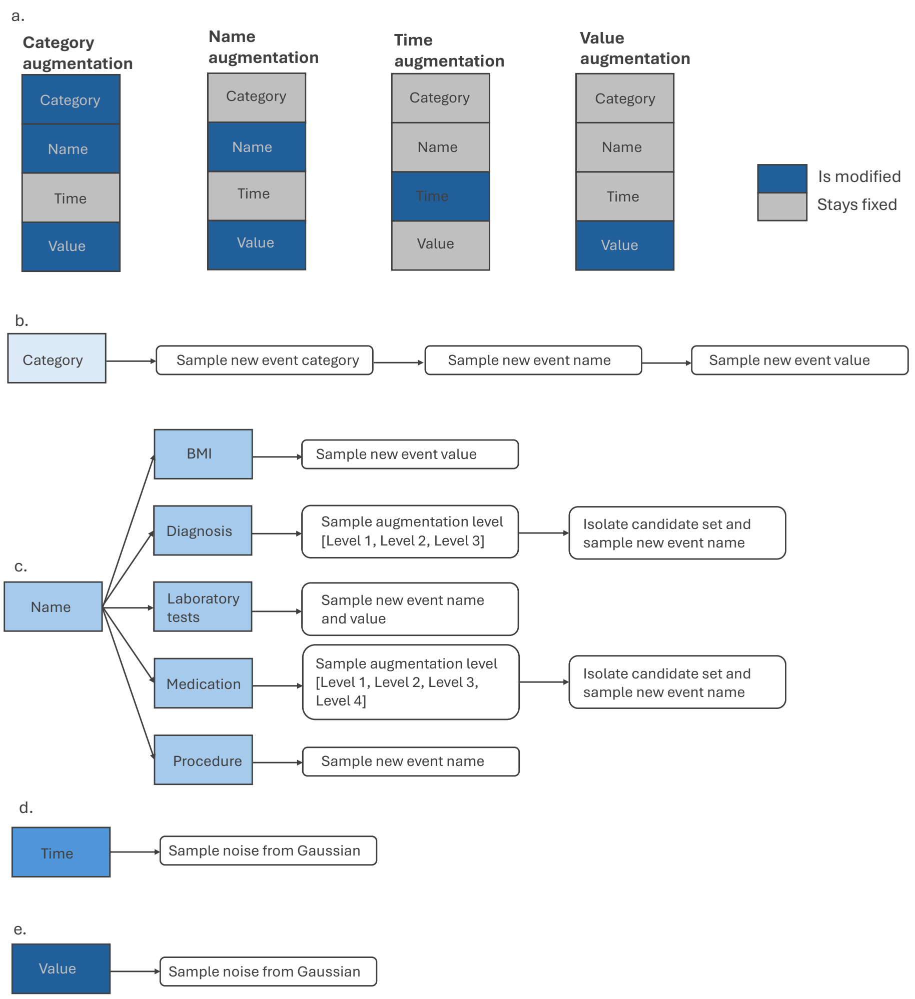


Supplementary Figure 3: The different augmentation types. (a) The event components that are modified in each augmentation type. (b) The steps in event category augmentation. (c) The steps in event name augmentation, depending on which event modality the name belongs to. (d) The steps in time noise augmentation. (e) The steps in event value augmentation.

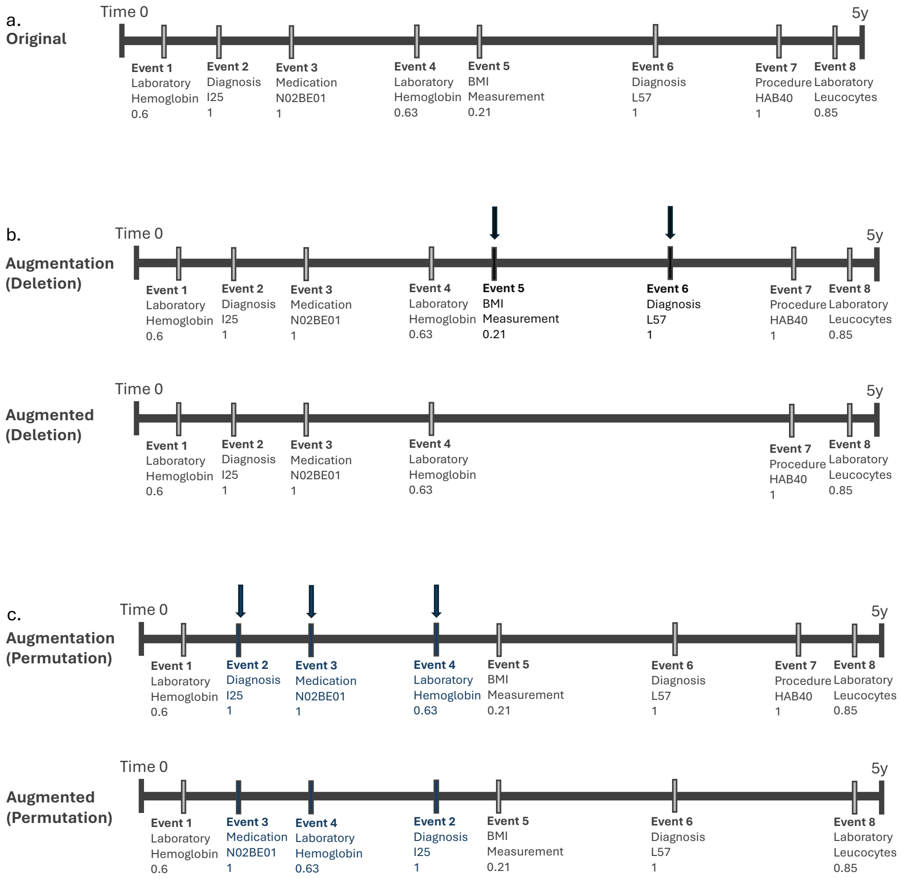


Supplementary Figure 4: Continues next page.

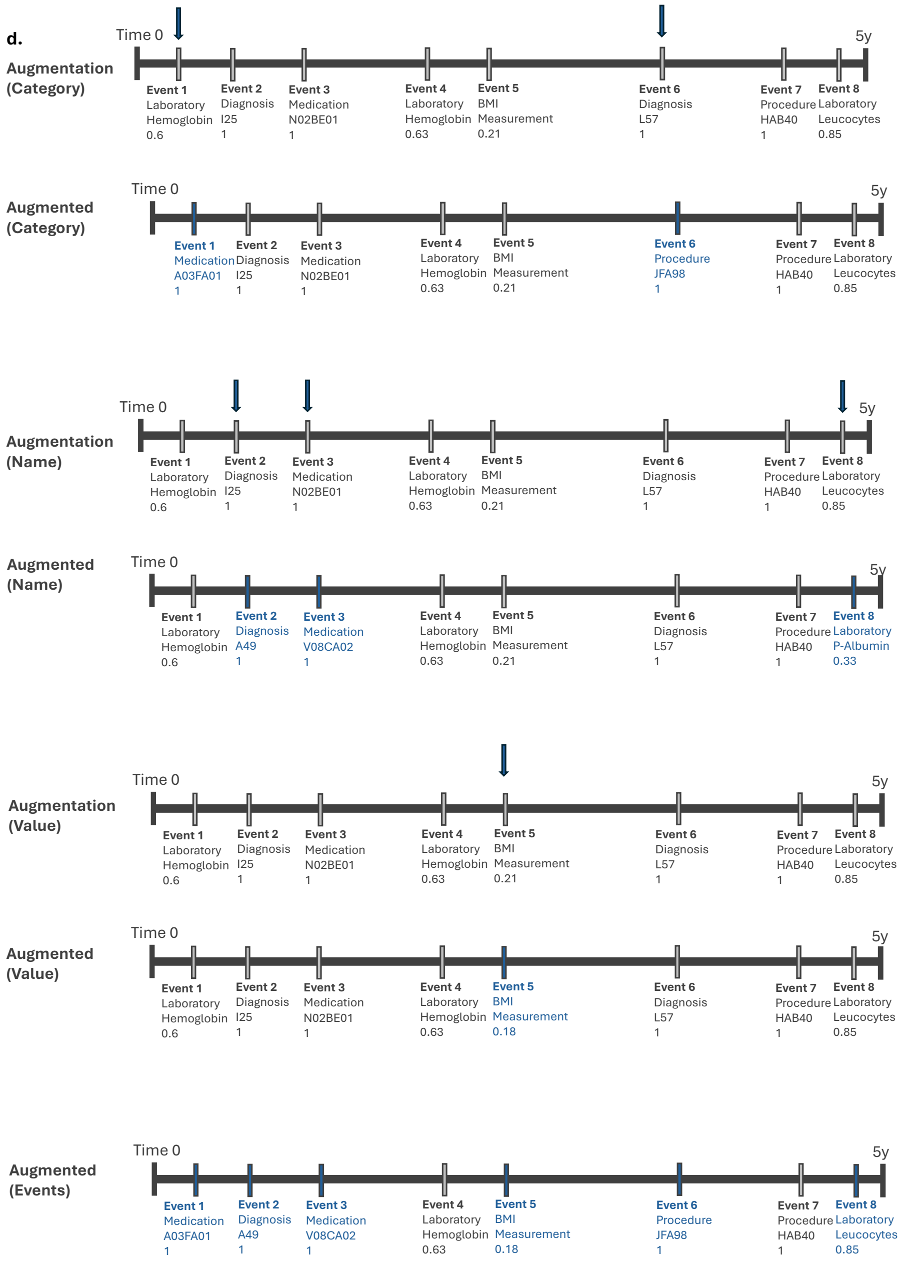


Supplementary Figure 4: Continue. Examples of stage one augmentations on event sequence. Time 0 is defined as the occurrence of the primary cancer diagnosis. (a) An example of an original event sequence. (b) Deletion of a window. (c) Permutation of a window. (d) Event-level augmentations. Category, name and value augmentations showed separately and finally as a combined result of the event-level augmentation.

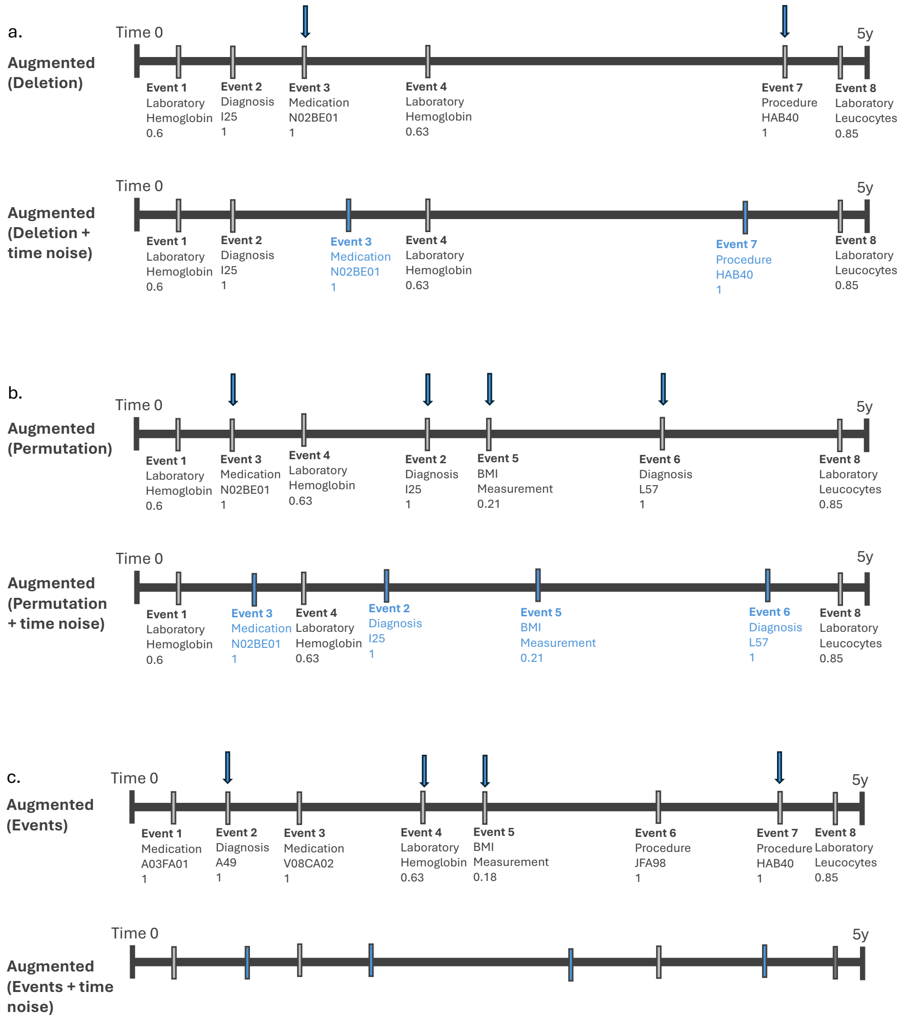


Supplementary Figure 5: Examples of stage two augmentations. (a) Time noise augmentation with deletion window. (b) Time noise augmentation with permutation window. (c). Time noise augmentation with event augmentations.

Supplementary Table 3: The used augmentation combinations. The chosen parameter values are bolded. The window length of extreme augmentations was sampled from [0.5, 0.8]. Numbers are probabilities unless otherwise mentioned. $p_{\text{deletion}}$ denotes the probability of event sequence having a window deleted and $p_{\text{permutation}}$ the probability of event sequence having a window permuted. $p_{\text{event}}$ denotes the probability of individual event being augmented, $p_{\text{time}}$ the probability of individual event getting noise added to their timestamp, and $\max_w$ the maximum number of weeks added to the timestamp.

| Combination | $p_{\text{deletion}}$ | $p_{\text{permutation}}$ | $p_{\text{event}}$ | $p_{\text{time}}$ | Noise range $[0, \max_w]$ |
|---|---|---|---|---|---|
| 1 | 0.1 | 0.1 | 0.2 | - | - |
| 2 | 0.1 | 0.1 | 0.3 | - | - |
| 3 | 0.1 | 0.1 | 0.5 | - | - |
| 4 | 0.2 | 0.2 | 0.2 | - | - |
| 5 | 0.2 | 0.2 | 0.3 | - | - |
| 6 | 0.2 | 0.2 | 0.5 | - | - |
| 7 | 0.1 | 0.1 | 0.3 | 0.1 | [0,2] |
| 8 | 0.1 | 0.1 | 0.3 | 0.5 | [0,2] |
| 9 | 0.1 | 0.1 | 0.3 | 1 | [0,2] |
| 10 | 0.1 | 0.1 | 0.3 | 0.1 | [0,12] |
| 11 | 0.1 | 0.1 | 0.3 | 0.5 | [0,12] |
| **12** | **0.1** | **0.1** | **0.3** | **1** | **[0,12]** |
| Extreme | 0.9 | 0.9 | 0.8 | 1 | [0,52] |

Supplementary Table 4: The categorical distribution from which the augmentation level of hierarchical diagnoses and medications was sampled. Probabilities for each level of hierarchy.

| | Level 1 | Level 2 | Level 3 | Level 4 |
|---|---|---|---|---|
| Diagnoses | 0.2 | 0.6 | 0.2 | - |
| Medications | 0.1 | 0.4 | 0.4 | 0.1 |

### 3 Event sequence embedding

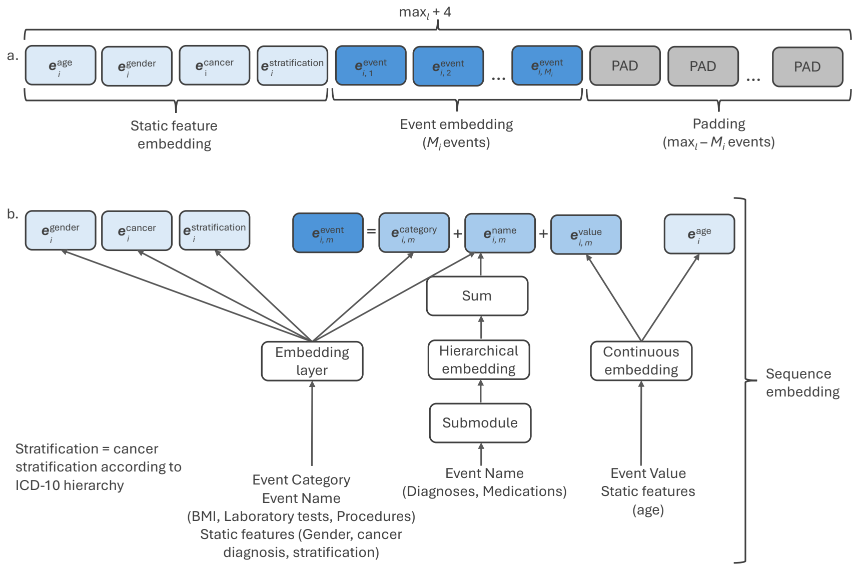


Supplementary Figure 6: (a) The structure of the event sequence of patient $i$. Index $i \in \{1, \ldots, N\}$. $M_i$ denotes the number of observed event of patient $i$ and $m \in \{1, \ldots, M_i\}$ indexes the events. Refer to Supplementary Table 1 for notations. (b). Embedding protocol for demographic information (light blue) and events (dark blue). The event embedding is the sum of category, name and value embeddings (blue). $\max_l$ is the maximum number of events in an event sequence across the dataset.

### 4 Submodules for diagnosis and medication code embeddings

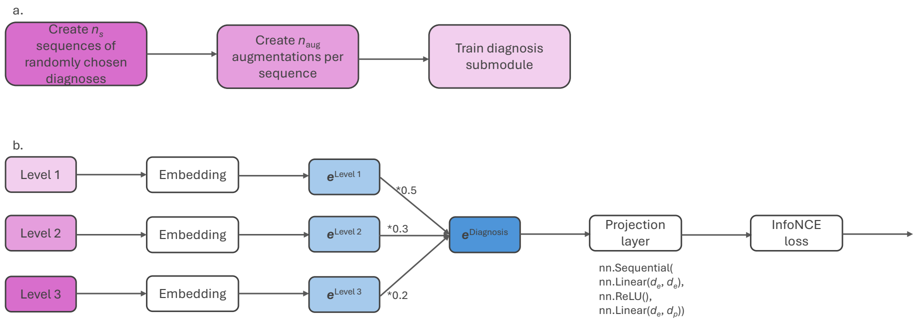


Supplementary Figure 7: The submodule for hierarchical embedding of diagnosis codes. Refer to Supplementary Table 1 for notations. (a) The steps involved in pre-training the diagnosis submodule. $n_s$ denotes the number of created sequences and $n_{\text{aug}}$ the number of created augmentations per sequence. (b) The diagnosis submodule architecture.

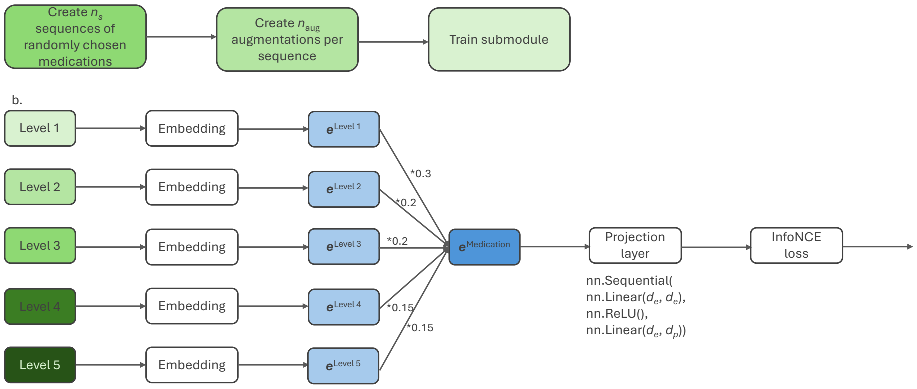


Supplementary Figure 8: The submodule for hierarchical embedding of medication codes. Refer to Supplementary Table 1 for notations. (a) The steps involved in pre-training the medication submodule. $n_s$ denotes the number of created sequences and $n_{\text{aug}}$ the number of created augmentations per sequence. (b) The medication submodule architecture.

### 5 Hyperparameter optimization

The model complexity, projection dimension, and embedding dimension were optimized in steps, freezing one parameter group at time (Supplementary Figure 9) with a simplified augmentation dataset. The optimization was performed with TACT. At each step, TACT was trained three times, using different random seeds and corresponding augmentation datasets. Parameters were selected based on the average performance across runs, evaluated with validation loss and event-level metrics. Dropout (0.2), InfoNCE temperature (0.5), learning rate ($10^{-0.3}$), and batch size (128) were kept fixed for computational efficiency

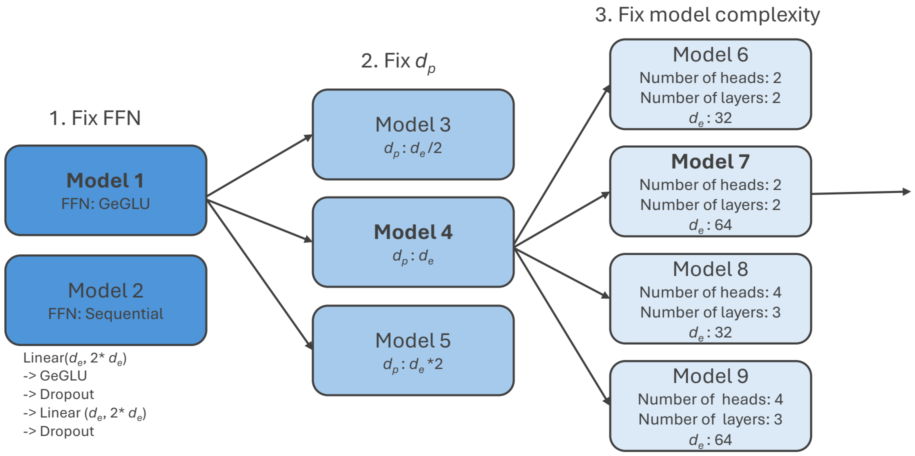


Supplementary Figure 9: The protocol of choosing model parameters. Chosen models in each stages are bolded. Refer to Supplementary Table 1 for notations.

A simplified augmentation dataset was constructed for hyperparameter optimization as follows:

1. Event category: for 15% of randomly chosen events, the event category was replaced with a token [MASK], used to encourage the model to learn relationships of event category, name, and value.

2. Event name: Applied only to diagnoses and medications. Hierarchy-based augmentation technique (described above) applied to all medication and diagnosis events.

3. Event value: Noise ($\mathcal{N} \sim (0, 0.05)$) was added for all laboratory measurements and BMI values.

4. Event time: Noise ($\mathcal{N} \sim (0, 2)$) was added for all events.

### 6 Final dataset

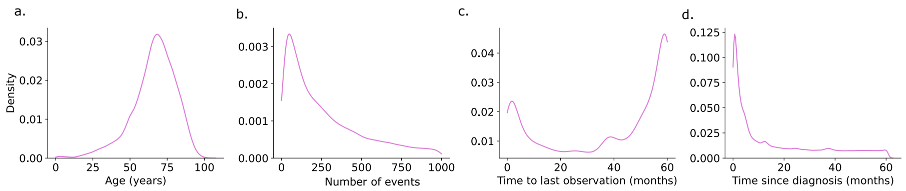


Supplementary Figure 10: The probability density function of (a) age at diagnosis. (b) number of events (sequence length) per patient. (c) length of follow-up period. (d) times to medical events in event sequences.

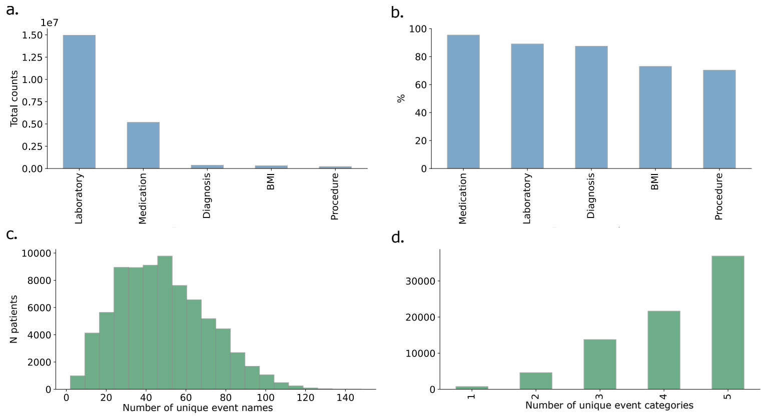


Supplementary Figure 11: (a) The abundance of data modalities. (b) The percentage of patients with at least one occurrence of a specific data modality. (c) The number of unique event names per patient. (d) The number of unique event categories per patient.

Supplementary Table 5: The 50 most abundant event names.

| Event name | Category | Total count | Number of patients with data |
|---|---|---|---|
| B -Hematocrit | Laboratory | 904847 | 68351 (88%) |
| B -Haemoglobin | Laboratory | 817104 | 68388 (88%) |
| B -Red blood cells | Laboratory | 803518 | 68289 (88%) |
| E -Mean Corpuscular Volume | Laboratory | 802422 | 68321 (88%) |
| E -Mean Corpuscular Haemoglobin Concentration | Laboratory | 801229 | 68225 (88%) |
| B -Platelets | Laboratory | 797717 | 68289 (88%) |
| B -White blood cells | Laboratory | 796759 | 68193 (88%) |
| E -Mean Corpuscular Haemoglobin | Laboratory | 795084 | 68186 (88%) |
| E -Red cell Distribution Width | Laboratory | 786176 | 68125 (88%) |
| P -Creatinine | Laboratory | 701901 | 66802 (86%) |
| P -Potassium | Laboratory | 661358 | 62063 (80%) |
| P -Sodium | Laboratory | 655227 | 61963 (80%) |
| Haemolysis index | Laboratory | 585381 | 61115 (79%) |
| P -C-reactive protein | Laboratory | 548784 | 60002 (77%) |
| N02BE01 | Medication | 409708 | 58463 (75%) |
| N02AA05 | Medication | 388677 | 47008 (61%) |
| P -Alanine aminotransferase | Laboratory | 315863 | 49234 (63%) |
| Measurement | (BMI) | 303435 | 56637 (73%) |
| B -Neutrophils | Laboratory | 297583 | 39592 (51%) |
| P -Alkaline phosphatase | Laboratory | 294910 | 48284 (62%) |
| Pt -Estimated glomerular filtration rate | Laboratory | 239970 | 35054 (45%) |
| B01AB05 | Medication | 209442 | 30098 (39%) |
| P -Bilirubin | Laboratory | 202069 | 36254 (47%) |
| P -Glucose | Laboratory | 178522 | 36662 (47%) |
| P -Calcium (total) | Laboratory | 172310 | 26568 (34%) |
| Lipemia index | Laboratory | 170039 | 32764 (42%) |
| Icterus index | Laboratory | 150623 | 26482 (34%) |
| P -Albumin | Laboratory | 146041 | 28101 (36%) |
| J01DC02 | Medication | 141817 | 25994 (33%) |
| P -Ionised calcium, actual | Laboratory | 129940 | 21609 (28%) |
| aB-Partial pressure of carbon dioxide | Laboratory | 127272 | 20724 (27%) |
| aB-pH | Laboratory | 127162 | 20714 (27%) |
| aB-Base excess | Laboratory | 125790 | 20335 (26%) |
| aB-Partial pressure of oxygen | Laboratory | 125345 | 20718 (27%) |
| aB-Standard bicarbonate | Laboratory | 123686 | 19910 (26%) |
| A02BC02 | Medication | 113743 | 32519 (42%) |
| fP-Plasma lactate | Laboratory | 112883 | 19175 (25%) |
| P -Ionised calcium | Laboratory | 112217 | 18743 (24%) |
| P -Prothrombin time | Laboratory | 108311 | 39172 (50%) |
| C03CA01 | Medication | 105852 | 20520 (26%) |
| V08AB02 | Medication | 100102 | 35391 (46%) |
| A04AA01 | Medication | 98588 | 34787 (45%) |
| P -Chloride | Laboratory | 95170 | 16300 (21%) |
| P -Prothrombin time, International normalized ratio | Laboratory | 93049 | 27560 (36%) |
| M01AE01 | Medication | 89453 | 28654 (37%) |
| B05BB01 | Medication | 89272 | 9290 (12%) |
| aB-Oxygen saturation, fraction of functional hemoglobin | Laboratory | 82669 | 13434 (17%) |
| C07AB07 | Medication | 81327 | 18369 (24%) |
| A12AX | Medication | 80906 | 21894 (28%) |
| aB-Methemoglobin | Laboratory | 76977 | 12339 (16%) |

### 7 Augmentation experiment details

Extreme augmentation experiments were performed on TACT. The model performance was evaluated at event-level and patient-level, similarly as when choosing the optimal augmentation combination.

Supplementary Table 8: The metrics on test data. The general, patient and event level metrics of extreme augmentations. Values reported as mean ± standard deviation (SD) across three runs unless otherwise specified. Here "mean", "min", and "max" refer to the average, minimum, and maximum value recorded for the corresponding metric. CAC % denotes the proportion of all available samples included in the neighborhood. Refer to Supplementary Table 1 for notations.

| Metric | Extreme augmentation |
|---|---|
| Val loss (↓) | 3.68 ± 0.01 |
| $DOR_{within\_patient}$ (mean, ↑) | 0.96 ± 0.01 |
| $DOR_{within\_patient}$ (min, ↑) | -0.057 ± 0.07 |
| $DOR_{within\_patient}$ (max, ↑) | 1.00 ± 0.00 |
| $DOR_{across\_patient}$ (mean, (↓) | 0.14 ± 0.01 |
| $DOR_{across\_patient}$(min, ↓) | -0.54 ± 0.07 |
| $DOR_{across\_patient}$ (max, ↓) | 1.00 ± 0.00 |
| $ADOR_{patient}$, ↑) | 0.45 ± 0.03 |
| Average cosine similarity of event categories (↓) | 0.25 ± 0.03 |
| $DOR_{within\_class}$ (↑) | 0.70 ± 0.01 |
| $DOR_{across\_class}$ (↓) | 0.06 ± 0.03 |
| $ADOR_{class}$ (↑) | 0.82 ± 0.02 |
| CAA (↑) | 0.74 ± 0.06 |
| CAC 5% (↑) | 0.98 ± 0.01 |
| CAC 50% (↑) | 0.69 ± 0.00 |

Supplementary Table 6: The metrics on validation data. The general, patient and event-level metrics of augmentation combinations. Values reported as mean ± standard deviation (SD) across three runs unless otherwise mentioned. Here "mean", "min", and "max" refer to the average, minimum, and maximum value recorded for the corresponding metric. CAC % denotes the proportion of all available samples included in the neighborhood. Refer to Supplementary Table 1 for notations.

| Aug comb | 1 | 2 | 3 | 4 | 5 | 6 |
|---|---|---|---|---|---|---|
| Val loss | 3.61 ± 0.00 | 3.61 ± 0.00 | 3.61 ± 0.00 | 3.61 ± 0.00 | 3.61 ± 0.00 | 3.61 ± 0.00 |
| $DOR_{within\_patient}$ (mean, ↑) | 0.99 ± 0.00 | 0.99 ± 0.00 | 0.99 ± 0.00 | 0.99±0.00 | 0.99 ± 0.00 | 0.99 ± 0.00 |
| $DOR_{within\_patient}$ (min, ↑) | 0.61 ± 0.20 | 0.63 ± 0.08 | 0.47 ± 0.07 | 0.33±0.10 | 0.24 ± 0.10 | 0.05 ± 0.05 |
| $DOR_{within\_patient}$ (max, ↑) | 1.00 ± 0.00 | 1.00 ± 0.00 | 1.00 ± 0.00 | 1.00 ± 0.00 | 1.00 ± 0.00 | 1.00 ± 0.00 |
| $DOR_{across\_patient}$ (mean, ↓) | 0.24 ± 0.01 | 0.22 ± 0.02 | 0.20 ± 0.01 | 0.24 ± 0.01 | 0.20 ± 0.01 | 0.21 ± 0.01 |
| $DOR_{across\_patient}$ (min, ↓) | -0.46 ± 0.05 | -0.50 ± 0.04 | -0.50 ± 0.03 | -0.44 ± 0.02 | -0.49 ± 0.02 | -0.45 ± 0.02 |
| $DOR_{across\_patient}$ (max, ↓) | 0.99 ± 0.00 | 0.99 ± 0.00 | 0.99 ± 0.00 | 0.99 ± 0.00 | 0.99 ± 0.00 | 0.99 ± 0.00 |
| $ADOR_{patient}$ (↑) | 0.90 ± 0.02 | 0.83 ± 0.04 | 0.70 ± 0.01 | 0.89 ± 0.01 | 0.81 ± 0.00 | 0.72 ± 0.02 |
| Average cosine similarity of event categories (↑) | 0.30 ±0.03 | 0.30 ± 0.02 | 0.36 ± 0.04 | 0.30 ±0.01 | 0.31 ± 0.01 | 0.32 ± 0.03 |
| $DOR_{within\_class}$ (↑) | 0.76 ± 0.02 | 0.76 ± 0.02 | 0.76 ± 0.02 | 0.76 ± 0.02 | 0.76 ± 0.02 | 0.76 ± 0.02 |
| $DOR_{across\_class}$ s (↓) | 0.12 ± 0.04 | 0.12 ± 0.03 | 0.19 ± 0.05 | 0.11 ± 0.01 | 0.13 ± 0.02 | 0.15 ± 0.04 |
| $ADOR_{class}$ (↑) | 0.88 ± 0.04 | 0.88 ± 0.03 | 0.83 ± 0.03 | 0.87 ± 0.03 | 0.87 ± 0.04 | 0.86 ± 0.04 |
| CAA (↑) | 0.76 ± 0.08 | 0.76 ± 0.09 | 0.69 ± 0.20 | 0.75 ± 0.09 | 0.75 ± 0.09 | 0.74 ± 0.10 |
| CAC 5% (↑) | 0.99 ± 0.03 | 0.99 ± 0.04 | 0.99 ± 0.07 | 0.99 ± 0.04 | 0.99 ± 0.04 | 0.99 ± 0.05 |
| CAC events 50% (↑) | 0.69 ± 0.03 | 0.69 ± 0.05 | 0.69 ± 0.06 | 0.69 ± 0.05 | 0.69 ± 0.04 | 0.69 ± 0.06 |

Supplementary Table 7: The metrics on validation data. The general, patient and event-level metrics of augmentation combinations. Values reported as mean ± standard deviation (SD) across three runs unless otherwise mentioned. Here, "mean", "min", and "max" refer to the average, minimum, and maximum value recorded for the corresponding metric. CAC % denotes the proportion of all available samples included in the neighborhood. Refer to Supplementary Table 1 for notations.

| Aug comb | 7 | 8 | 9 | 10 | 11 | 12 |
|---|---|---|---|---|---|---|
| Val loss | 3.61 ± 0.00 | 3.61 ± 0.00 | 3.63 ± 0.03 | 3.61 ± 0.00 | 3.61 ± 0.00 | 3.62 ± 0.00 |
| $DOR_{within\_patient}$ (mean, ↑) | 0.99 ± 0.00 | 0.99 ± 0.00 | 0.99 ± 0.00 | 0.99 ± 0.00 | 0.99 ± 0.00 | 0.99 ± 0.00 |
| $DOR_{within\_patient}$ (min, ↑) | 0.65 ± 0.10 | 0.72 ± 0.05 | 0.71 ± 0.10 | 0.69 ± 0.04 | 0.68 ± 0.09 | 0.80 ± 0.05 |
| $DOR_{within\_patient}$ (max, ↑) | 1.00 ± 0.00 | 0.99 ± 0.00 | 0.99 ± 0.00 | 1.00 ± 0.00 | 0.99 ± 0.00 | 0.99 ± 0.00 |
| $DOR_{across\_patient}$ (mean, ↓) | 0.20 ± 0.00 | 0.22 ± 0.03 | 0.20 ± 0.02 | 0.21 ± 0.02 | 0.19 ± 0.02 | 0.19 ± 0.01 |
| $DOR_{across_patient}$ (min, ↓) | -0.47 ± 0.02 | -0.47 ± 0.06 | -0.52 ± 0.03 | -0.50 ± 0.00 | -0.53 ± 0.01 | -0.48 ± 0.05 |
| $DOR_{across\_patient}$ (max, ↓) | 0.99 ± 0.00 | 0.99 ± 0.00 | 0.99 ± 0.00 | 0.99 ± 0.00 | 0.99 ± 0.00 | 0.99 ± 0.00 |
| $ADOR_{patient}$ (↑) | 0.83 ± 0.01 | 0.87 ± 0.01 | 0.86 ± 0.02 | 0.85 ± 0.01 | 0.87 ± 0.01 | 0.84 ± 0.02 |
| Average cosine similarity of event categories (↑) | 0.32 ± 0.31 | 0.28 ±0.03 | 0.28 ± 0.04 | 0.29 ± 0.04 | 0.27 ± 0.03 | 0.25 ± 0.01 |
| $DOR_{within\_class}$ (↑) | 0.76 ± 0.03 | 0.75 ± 0.02 | 0.74 ±0.01 | 0.75 ± 0.02 | 0.73 ± 0.02 | 0.74 ± 0.01 |
| $DOR_{across\_class}$ (↓) | 0.14 ± 0.05 | 0.09 ± 0.03 | 0.09 ± 0.04 | 0.11 ± 0.05 | 0.08 ± 0.04 | 0.06 ± 0.01 |
| $ADOR_{class}$ (↑) | 0.86 ± 0.04 | 0.87 ± 0.02 | 0.88 ± 0.03 | 0.87 ± 0.03 | 0.88 ± 0.04 | 0.89 ± 0.02 |
| CAA (↑) | 0.74 ± 0.10 | 0.75 ± 0.07 | 0.77 ± 0.06 | 0.76 ± 0.08 | 0.75 ± 0.08 | 0.77 ± 0.05 |
| CAC 5% (↑) | 0.99 ± 0.04 | 0.99 ± 0.05 | 0.99 ± 0.07 | 0.99 ± 0.04 | 0.99 ± 0.08 | 0.99 ± 0.06 |
| CAC 50% (↑) | 0.69 ± 0.07 | 0.69 ± 0.05 | 0.69 ± 0.03 | 0.69 ± 0.05 | 0.69 ± 0.06 | 0.69 ± 0.04 |

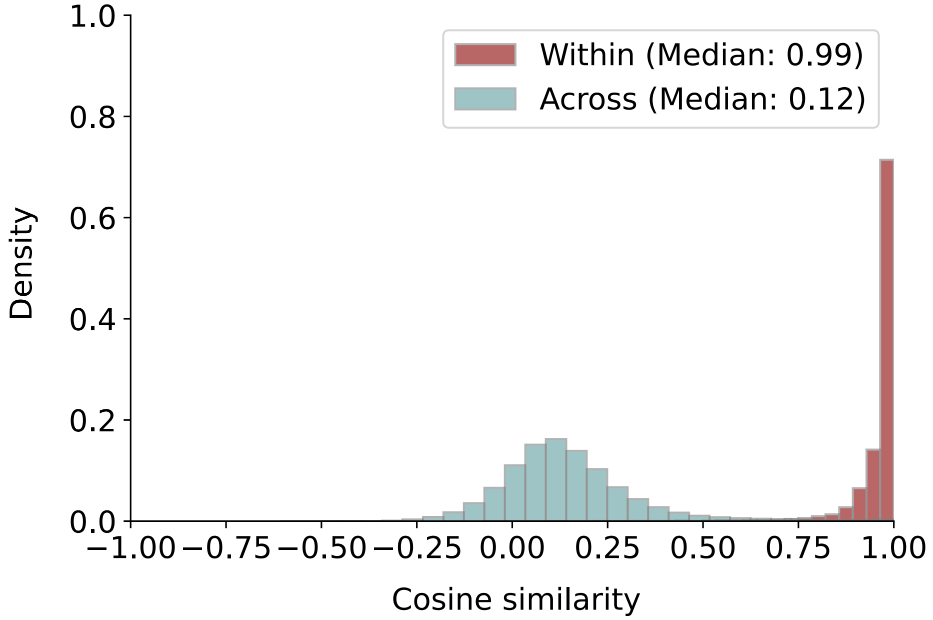


Supplementary Figure 12: Cosine similarities of extreme augmentations, TACT. Within (original-augmented pair) and across (non-pairs) cosine similarities.

### 8 Event-level Model performances

Supplementary Table 9: The metrics on test data. Category specific metrics across three runs. The best performance is bolded. Values reported as mean ± standard deviation (SD) unless otherwise mentioned. BMI was excluded from further analysis for having only one event. Refer to Supplementary Table 1 for notations.

| | TACT | TACT without pre-training | ClinicalTAAT |
|---|---|---|---|
| $DOR_{within\_class}$ (↑) | | | |
| Diagnoses | **0.76 ± 0.03** | 0.58 ± 0.04 | 0.59 ± 0.02 |
| Laboratory tests | 0.39 ± 0.04 | 0.42 ± 0.03 | **0.47 ± 0.02** |
| Medications | **0.82 ± 0.01** | 0.49 ± 0.02 | 0.54 ± 0.02 |
| Procedures | 0.55 ± 0.03 | 0.52 ± 0.03 | **0.57 ± 0.02** |
| $DOR_{across\_class}$ (↓) | | | |
| Diagnoses | 0.07 ± 0.05 | 0.05 ± 0.04 | **-0.04 ± 0.02** |
| Labs | 0.03 ± 0.08 | 0.04 ± 0.02 | **0.01 ± 0.05** |
| Medications | 0.08 ± 0.03 | 0.06 ± 0.01 | **-0.01 ± 0.01** |
| Procedures | 0.28 ± 0.32 | 0.09 ± 0.04 | -0.01 ± 0.04 |
| $ADOR_{class}$ (↑) | | | |
| Diagnoses | **1.00 ± 0.00** | 0.53 ± 0.32 | 0.91 ± 0.08 |
| Labs | 0.09 ± 0.13 | 0.07 ± 0.11 | **0.32 ± 0.29** |
| Medications | **1.00 ± 0.00** | 0.04 ± 0.03 | 0.52 ± 0.14 |
| Procedures | 0.52 ± 0.13 | 0.07 ± 0.02 | 0.76 ± 0.08 |
| CAA(↑) | | | |
| Diagnoses | **0.87 ± 0.23** | 0.09 ± 0.08 | 0.51 ± 0.16 |
| Labs | 0.01 ± 0.02 | 0.03 ± 0.01 | **0.28 ± 0.16** |
| Medications | **1.00 ± 0.00** | 0.02 ± 0.02 | 0.29 ± 0.13 |
| Procedures | 0.04 ± 0.05 | 0.02 ± 0.02 | **0.43 ± 0.04** |
| CAC **5%** (↑) | | | |
| Diagnoses | 1.00 ± 0.00 | 1.00 ± 0.00 | 1.00 ± 0.00 |
| Labs | 0.71 ± 0.20 | 0.90 ± 0.03 | **0.95 ± 0.02** |
| Medications | 1.00 ± 0.00 | 1.00 ± 0.00 | 1.00 ± 0.00 |
| Procedures | 0.99 ± 0.00 | 0.99 ± 0.00 | **1.00 ± 0.00** |
| CAC **50%** (↑) | | | |
| Diagnoses | 0.99 ± 0.00 | 0.99 ± 0.00 | 0.99 ± 0.00 |
| Labs | 0.71 ± 0.2 | 0.90 ± 0.03 | 0.95 ± 0.02 |
| Medications | 0.99 ± 0.00 | 0.98 ± 0.01 | 0.99 ± 0.00 |
| Procedures | 0.91 ± 0.03 | 0.96 ± 0.01 | 0.99 ± 0.00 |
